# Network-based spreading of grey matter changes across different stages of psychosis

**DOI:** 10.1101/2022.01.11.22268989

**Authors:** Sidhant Chopra, Ashlea Segal, Stuart Oldham, Alexander Holmes, Kristina Sabaroedin, Edwina R. Orchard, Shona M. Francey, Brian O’Donoghue, Vanessa Cropley, Barnaby Nelson, Jessica Graham, Lara Baldwin, Jeggan Tiego, Hok Pan Yuen, Kelly Allott, Mario Alvarez-Jimenez, Susy Harrigan, Ben D. Fulcher, Kevin Aquino, Christos Pantelis, Stephen J Wood, Mark Bellgrove, Patrick McGorry, Alex Fornito

## Abstract

**Importance:** Psychotic illness is associated with anatomically distributed grey matter reductions that can worsen with illness progression, but the mechanisms underlying the specific spatial patterning of these changes is unknown.

**Objective:** To test the hypothesis that brain network architecture constrains cross-sectional and longitudinal grey matter alterations across different stages of psychotic illness and to identify whether certain brain regions act as putative epicentres from which volume loss spreads.

**Design, Settings, Participants:** This study included 534 individuals from 4 cohorts, spanning early and late stages of psychotic illness. Early-stage cohorts included patients with antipsychotic-naïve first episode psychosis (N=59) and a group of medicated patients within 3 years of psychosis onset (N=121). Late-stage cohorts comprised two independent samples of people with established schizophrenia (N=136 in total). Each patient group had a corresponding matched control group (N=218 in total). A further independent sample of healthy adults (N=346) was used to derive representative structural and functional brain networks for modelling of network-based spreading processes. We additionally examined longitudinal illness-related and antipsychotic-related grey matter changes over 3 and 12 months using a triple-blind randomised placebo-control MRI study of the antipsychotic-naïve patients. All data were collected between April 2008 and January 2020, and analyses were performed between March 2021 and January 2023.

**Main Outcomes and Measures:** We used coordinated deformation models to predict the extent of grey matter volume change in each of 332 parcellated areas by the volume changes observed in areas to which they were structurally or functionally coupled. To identify putative epicentres of volume loss, we used a network diffusion model to simulate the spread of pathology from different seed regions. Correlations between predicted and empirical spatial patterns of grey matter volume alterations were used to quantify model performance.

**Results:** In both early and late stages of illness, spatial patterns of cross-sectional volume differences between patients and controls were more accurately predicted by coordinated deformation models constrained by structural, rather than functional, network architecture (. 46 < *r* < .57; p < .001). The same model also robustly predicted longitudinal volume changes related to illness (*r* > 52; *p* < .001) and antipsychotic exposure (*r* > .50; *p* < .001). Diffusion modelling consistently identified, across all four datasets, the anterior hippocampus as a putative epicentre of pathological spread in psychosis (*all p* < .05). Epicentres of longitudinal grey matter loss were apparent posteriorly early in the illness and shifted anteriorly to prefrontal cortex with illness progression.

**Conclusion and Relevance:** Our findings highlight a robust and central role for white matter fibres as conduits for the spread of pathology across different stages of psychotic illness, mirroring findings reported in neurodegenerative conditions. The structural connectome thus represents a fundamental constraint on brain changes in psychosis, regardless of whether these changes are caused by illness or medication. Moreover, the anterior hippocampus represents a putative epicentre of early brain pathology from which dysfunction may spread to affect connected areas.

**Key points:** *Question:* Are grey matter changes across the psychosis continuum constrained by brain network architecture and are certain regions epicentres of volume loss?

*Findings:* Across four independent samples spanning different stages of psychotic illness, grey matter alterations are strongly constrained by the underlying architecture of the brain’s axonal pathways and the hippocampus is consistently identified as a putative source from which volume-loss may spread to connected regions.

*Meaning:* White matter fibres may act as conduits for the spread of pathology across all stages of psychotic illness and medial temporal regions play a critical role in the origins of grey matter reductions.

## Introduction

Psychotic disorders such as schizophrenia are characterised by anatomically distributed reductions in grey-matter volume (GMV)^1-7^, many of which show evidence of progression over time and across different stages of illness^8-14^. Meta- and mega-analyses indicate that some of the most robust cross-sectional GMV reductions are found in frontal, cingulate and temporal cortices, as well as medial temporal lobe and thalamus^3-7,15-17^, with longitudinal reductions identified in frontal, temporal and parietal cortices^9,11^. However, despite a large literature describing the location and nature of these brain changes, the specific mechanisms that give rise to their characteristic spatial pattern remain unknown.

The human brain is an intricate network of functionally specialised regions linked by a complex web of axonal fibres, referred to as a connectome^18^. These fibres enable the widespread coordination of neuronal dynamics and the transport of trophic and other biological molecules throughout the brain. They can also act as conduits for the spread of pathology, such that illness processes originating in one area can propagate to affect distributed systems via multiple mechanisms^19,20^. This principle has been powerfully demonstrated in dementia, where GMV reductions in different neurodegenerative conditions have been shown to spread through the brain in a way that is constrained by the underlying architecture of the brain’s white-matter pathways^21-24^.

Recent work suggests that a network-based spreading process may also be involved in psychosis. Cross-sectional grey-matter reductions in patients correlate with increased fractional anisotropy in regionally adjacent white matter^25^, are often correlated across spatially distributed regions^26-28^, and correspond with normative connectome organisation^29,30^. In recent work, Shafiei, et al. ^31^ developed a coordinated-deformation model (CDM) in a sample of people with established schizophrenia that predicted the level of cross-sectional GMV reduction in an area based on the average reductions observed in other areas to which it was structurally connected.

Together, these findings support the hypothesis that the spatial patterning of GMV loss in psychotic illness is constrained by connectome architecture. However, the few studies addressing this question have been cross-sectional and only examined patients with chronic illness, precluding an opportunity to track how coordinated grey-matter changes evolve through time and across illness stages. It thus remains unclear whether longitudinal GMV changes are actually constrained by brain network architecture, as would be expected for a network-based spreading process. Moreover, the reliance on samples of patients taking antipsychotic medication makes it difficult to determine whether coupled grey-matter changes are driven by treatments for the illness or the illness process itself.

Here, we used multiple cohorts spanning different stages of psychosis to comprehensively evaluate network constraints on cross-sectional and longitudinal GMV changes. Specifically, we evaluated the capacity of different CDM variants, constrained by distinct aspects of connectome structure or function, to model cross-sectional GMV differences in two samples of patients at early illness stages and two samples of patients at later stages, allowing for independent replication of our findings at each stage. The early-stage samples comprised a group of antipsychotic-naïve first episode psychosis patients and a group of medicated patients within 3 years of illness onset. The late-stage samples comprised two independent samples of people with established schizophrenia. We additionally leveraged longitudinal data acquired within the context of a longitudinal, randomised placebo-controlled study in the antipsychotic-naïve FEP group^32,33^ to evaluate the degree to which the CDMs predicted longitudinal GMV changes attributable to either antipsychotic medication or the illness process itself, as observed over 3- and 12-month intervals. We then used a network-based diffusion model to simulate the dynamic progression of GMV loss from different seed areas to determine whether specific brain regions act as putative sources or epicentres of network-based spread. This approach thus allowed us to robustly investigate the degree to which brain network architecture constrains a diverse array of cross-sectional and longitudinal GMV pathology across different stages of psychosis and to identify possible focal points of early brain volume loss.

## Methods

### Sample characteristics

This study used data from four independent datasets sampling different stages of the psychotic-illness continuum: the STAGES clinical trial^13,33^ (first episode psychosis; FEP), Human Connectome Project Early Psychosis^34^ (early psychosis; EP), BrainGluSchi^35^ (schizophrenia; SCZ-BGS), and COBRE^36^ (schizophrenia; SCZ-COBRE). Hereafter, these cohorts will be referred to as FEP, EP, SCZ-BGS and SCZ-COBRE, respectively. We also derived representative structural and functional connectomes using a large independent healthy control sample. The final number of included subjects, demographic and diagnostic characteristics are described in Table 1 and additional details about each dataset are provided in the Supplement1A.

**Table 1.**
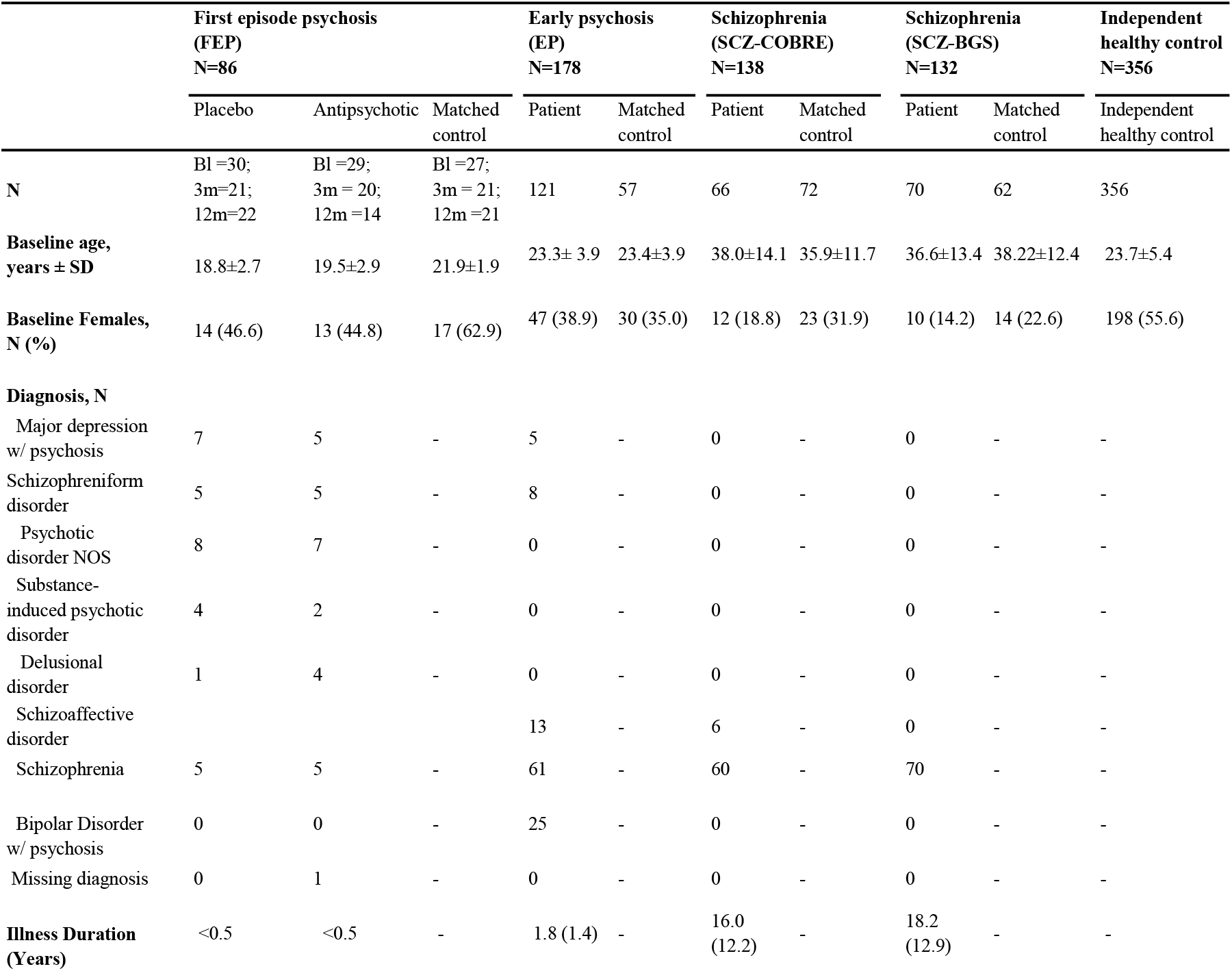
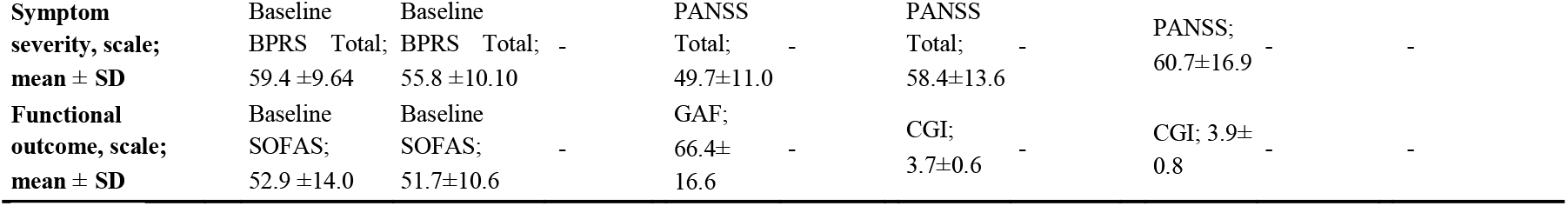
Sample characteristics of included datasets. Abbreviations: NOS = not otherwise specified; BPRS = Brief Psychiatric Rating Scale version 4; SOFAS = Social and Occupational Functioning Assessment Scale; Bl = baseline, 3m = 3-months, 12m = 12months; PANSS = Positive and Negative Syndrome Scale, GAF = Global Assessment of Functioning Scale (average of the social and occupational rating items); CGI: The Clinical Global Impressions Scale.

#### Structural MRI processing

Acquisition parameters for structural MRI can be found in Supplement1B. Prior to processing, raw T1w scans were visually examined for artefacts and then subjected to an automated quality control procedure^37^. In the FEP, EP, SCZ-BGS and SCZ-COBRE datasets, three, eight, six and four patient scans did not pass image quality control, respectively, and were excluded due to artefacts (see Supplement). The remaining scans were processed using the deformation-based morphometry (DBM) pipeline of the Computational Anatomy Toolbox (version r1113)^38^ for the Statistical Parametric Mapping 12^39^ software running in MATLAB version 2019a (details in the Supplement1C). We used DBM to quantify volume changes because it does not require tissue segmentation, requires less spatial smoothing^40^ than voxel-based morphometry (VBM) and to be comparable to previous work^24,31^. However, we replicated our primary findings using VBM (see *Robustness analyses*).

#### Quantifying cross-sectional and longitudinal grey matter changes in patients

To map spatial patterns of group-level cross-sectional and longitudinal volume change, we used a robust marginal model implemented in the Sandwich Estimator Toolbox^41^, which combines ordinary least squares estimates of parameters of interest with estimates of variance/covariance based on a robust sandwich estimator, thus accounting for within-subject correlations in longitudinal studies. This method is asymptotically robust to misspecification of the covariance model and does not depend on the assumptions of common longitudinal variance structure across the whole brain. All contrasts were adjusted for age, sex, and handedness, with site additionally included for the EP dataset.

We conducted cross-sectional contrasts in each of the four patient datasets to capture cross-sectional GMV differences between patients and controls (Fig1A). Longitudinal GMV changes were mapped in the FEP dataset (Fig1A) to isolate: (1) illness-related change over time, by comparing GMV changes overt time in the placebo group to matched healthy controls; and (2) antipsychotic-related changes over time, which compared GMV changes in the medication group to both the placebo group and matched healthy controls (see also^13^). Longitudinal contrasts were assessed from baseline to 3 months and baseline to 12 months, with a linear contrast being used for the latter. Contrasts were specified such that positive values in the resulting voxel-wise *t*-statistic maps indicate lower volume in patients compared to controls at cross-sectional contrast, and a greater longitudinal GMV decline in patients compared to controls in the longitudinal contrasts. The *t*-statistics were converted to z-scores, and we applied the CDM to unthresholded *z*-maps encoding regional GMV changes, as we are interested in capturing the spatial patterning of GMV differences across the entire brain, not just the changes which survive a statistical threshold. Renderings of the unthresholded *t*-maps can be found in Fig1A-C and Fig2A-B. FDR-corrected and uncorrected voxel-level *t*-statistic maps for each contrast are provided in the Supplements1F-1G.

**Fig 1.**
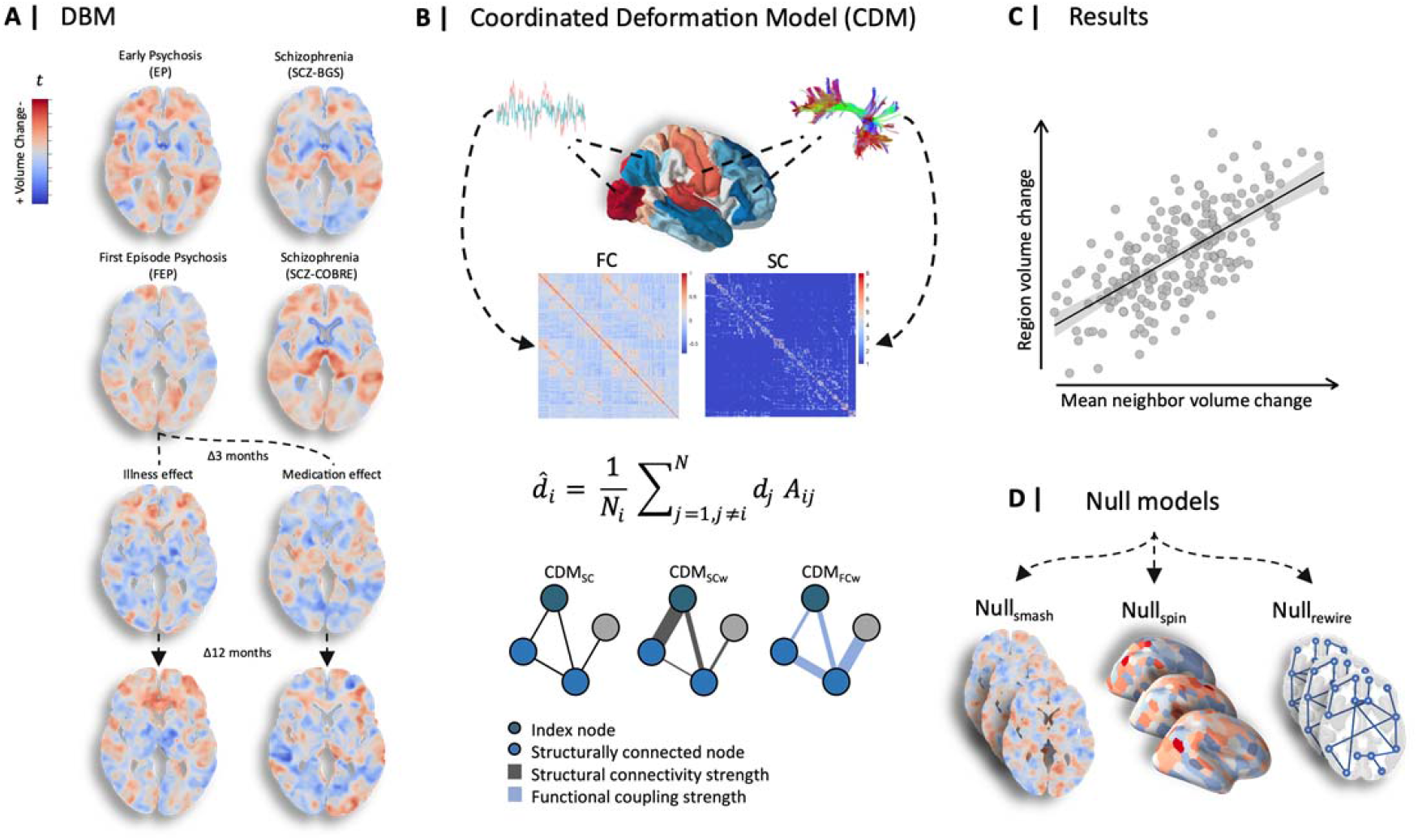
Analysis workflow for the Coordinated Deformation Model. (A) We derived voxel-wise GMV estimates using Deformation-based morphometry (DBM). Five separate contrasts were specified using a robust marginal model to infer baseline GMV differences and longitudinal GMV changes associated with illness and antipsychotic medication at 3 months and 12 months. (B) The contrast statistics were mapped to a brain parcellation comprising 332 regions, and diffusion and functional MRI data from an independent healthy sample were used to generate sample-averaged functional coupling (FC) and structural connectivity (SC) matrices. These matrices were used to model average volume changes in structurally connected neighbours. Under the CDM, the predicted deformation of a node, 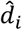, is modelled as a weighted sum of the deformation values observed its structurally connected neighbours (shown as light blue nodes in the example graphs). The weights are given by the adjacency matrix, *A*_*ij*_. Three different matrices were used, yielding three CDM variants; (1) A model denoted as CDM_SC,_ in which *A*_*ij*_ = 1 if two regions share a connection and *A*_*ij*_ = 0 otherwise; (2) a model denoted as CDM_SCw_ in which the elements of *A*_*ij*_ correspond precisely to the weighted SC matrix, such that the contribution of each neighbour is weighted by the strength of its structural connectivity to the index node; and (3) a model denoted CDM_FCw,_ in which the elements of *A*_*ij*_ correspond precisely to the weighted FC matrix, such that the contribution of each neighbour is weighted by its FC with the index node. (C) Model performance was evaluated using the Pearson correlation between regional estimates of observed and predicted GMV differences. (D) We also compared model performance to three benchmark null models accounting for spatial autocorrelations in the deformation maps (*Null*_*smash*_ and *Null*_*spin*_) and basic topological properties of the connectome (*Null*_*rewire*_; see *CDM evaluation)*.

**Fig 2.**
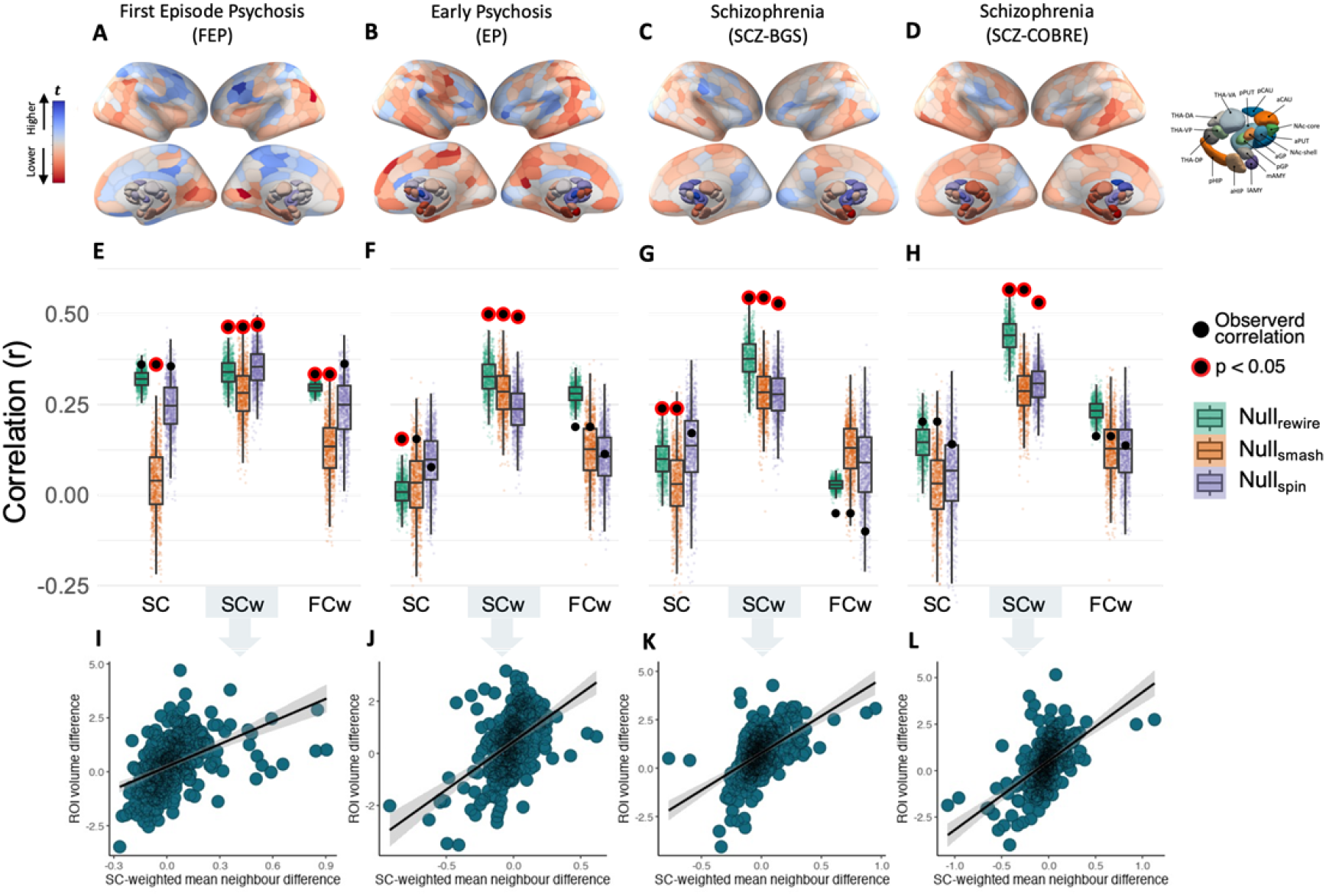
Baseline and longitudinal illness-related GMV changes are constrained by connectome anatomy. **(A-D)** The contrast statistics for four cross-sectional contrasts mapped to a brain parcellation comprising 332 regions. **(E-H)** Performance of the equally weighted (CDM_SC_), structural connectivity-weighted (CDM_SCw_), and functional coupling-weighted (CDM_FCw_) models relative to the Null_smash_, Null_spin_, and Null_rewire_ benchmarks. Black circles indicate the observed rank correlations between predicted and actual regional deformation values for each model at each timepoint, with red borders indicating statistical significance. Note that the observed value used for comparison against the Null_spin_ models is different because the subcortex was excluded. **(I–L)** Scatterplots of the association between observed and predicted regional volume deformation values for the best performing CDM_SCw_ model at each timepoint.

#### Brain Parcellation

To relate grey-matter alterations to connectome architecture, we parcellated the brain into 300 discrete cortical regions of approximately equal size^42^, in addition to 32 subcortical areas^43^, using previously validated atlases. The volume change for each region was estimated as the mean *z-*statistic of all voxels corresponding to that region. The regions comprise the nodes of a network, which can then be directly related to measures of inter-regional SC and FC.

#### Healthy reference connectomes

We derived a group-level healthy structural connectome from diffusion-weighted imaging (DWI) data from an independent sample of 356 adults (Fig1B; Table 1), which served as a reference connectome for computational modelling. Acquisition parameters and detailed overview of DWI processing and optimisation can be found in Supplement1D. This procedure resulted in a single 332 × 332 weighted group-average SC matrix.

We also derived a group-level healthy functional connectome from resting-state fMRI data acquired in the same independent sample of adults (Fig1B). Acquisition parameters for functional MRI and detailed information on fMRI processing can be found in Supplement1E. Given ongoing controversy around the application of global signal regression^44,45^, we evaluated how this step affected our findings (see *Robustness analyses*). After processing and denoising, we computed a whole-brain 332 × 332 FC matrix for each subject using pair-wise Pearson correlations between the timeseries of each of the 332 regions and finally took a mean FC matrix across the sample.

#### Coordinated Deformation Model (CDM) – Network Constraints

We evaluated network constraints on cross-sectional and longitudinal GMV changes using the CDM introduced by Shafiei, et al. ^31^. The model is given by

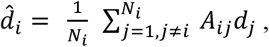

where 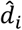 is the predicted GMV change in node *i, N*_*i*_ is the number of structurally connected neighbours of *i, d*_*j*_ is the deformation observed in the *j*-th neighbour of node *i*, and *A*_*ij*_ defines the connectivity between nodes *i* and *j*.

Three different matrices were substituted for *A*_*ij*_, yielding three variants of the CDM (Fig1B). For the first model, denoted CDM_SC_, *A*_*ij*_=1 if nodes *i* and *j* are connected in the group-average SC matrix and zero otherwise. Therefore, all *j* structurally connected neighbours make an equal contribution to predicting the extent of deformation observed in node *i*.

For the second and third models, denoted CDM_SCw_ and CDM_FCw_, *A*_*ij*_ corresponded to the weighted SC or FC matrices, respectively. Therefore, under these models, the contributions of node *i*’s neighbours were weighted by either inter-regional SC (CDM_SCw_) or FC (CDM_FCw_) estimates, such that neighbours of node *i* with a more strongly weighted connection made a stronger contribution to predicting node *i*’s volume change (Fig1B). In all models, only edges that had a corresponding structural connection were included and SC and FC edge weights were taken from the healthy reference connectome, unless otherwise specified.

#### CDM evaluation

Model performance was evaluated using the product-moment correlation (*r*) between region-wise estimates of observed and predicted deformation (Fig1C). We also compared the performance of the CDM_SC_, CDM_SCw_ and CDM_FCw_ models to three benchmark null models. The first (Fig1D; Null_smash_) and second (Fig1D; Null_spin_) null models evaluated whether the observed findings were specific to the empirically observed pattern of grey-matter deformations or were a generic property of the intrinsic spatial correlation structure of the deformation maps. The third null model (Fig1D; Null_rewire_) tests the hypothesis that any network-based prediction of local grey-matter change is specific to the actual topology of the connectome itself and cannot be explained by basic network properties, such as regional variations in node degree or the spatial dependence of inter-regional connectivity^46^. Further details about benchmark null models can be found in the Supplement1H.

#### Network Diffusion Model (NDM) – Epicentre Identification

The CDM evaluates the degree to which spatial patterns of GMV change are shaped by connectome properties. A close coupling between GMV change and network architecture implies that volume loss spreads through the connectome, but the CDM offers limited insight into the dynamics of the spreading process, nor is it able to identify regions from which the spreading may initiate. We therefore used a NDM to directly test whether GMV loss spreads through the brain via a process of diffusion and whether certain brain regions act as sources, or epicentres, of pathological spread through the brain (Fig4)^47^. The NDM simulates the dynamic spread of pathology between the nodes of a weighted network via a process of diffusion (Fig4A), defined as

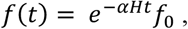

where *t* is the model diffusion time, which has arbitrary units (a.u.), and *f*(*t*) is a vector characterizing the amount of diffusion in each region at time *t*. The strength of the diffusion process is controlled by a constant (*α*) and *H* is the Laplacian of the weighted SC matrix. *f*_0_ represents the initial distribution of pathology. We repeatedly initialized the model using each of the 332 regions as the starting seed, such that the initial state was set to 1 for the seed region, and 0 for all other regions. At each initialization, using a constant of *α* = 1, the NDM was used to estimate the diffusion at all other regions at time *t* = 0 to 50. In this way, we were able to determine whether a diffusion process seeded from each region resulted in a spatial distribution of volume loss that matched the empirically observed patterns. Further information about the NDM can be found in the Supplement1I.

#### NDM evaluation

Consistent with prior work ^47,48^, model performance was evaluated as the Pearson correlation between the predicted diffusion and observed volume abnormalities at each time step and for each seed, with the maximum correlation (Fig4A; *r*_*max*_) across all time steps being retained. The observed regional *t*-statistics were rescaled to a more interpretable non-negative quantity via a log-transformation, yielding values in the range [0,1]^47-49^. The seed region was excluded when correlating predicted and observed volume abnormalities to ensure that our analysis was not influenced by large volume abnormalities in the seeds. The performance of the NDM in capturing the empirical maps of GMV change was compared to its performance in capturing surrogate maps generated using the Null_smash_ and Null_rewire_ benchmark models (Fig4A). The Null_spin_ benchmark was not used to evaluate the NDM as it does not include subcortical regions. Further details about benchmark null models used to evaluate the NDM can be found in the Supplement1I.

To aid comparison with previous research^31^, we also implemented a data-driven approach to epicentre identification that defines epicentres as regions with high volume loss that are also connected to other regions with high volume loss (FigS2; see Supplement1J for details). The spatial locations of epicentres identified by this approach closely aligned with the results of the NDM epicentre analysis.

## Results

### Structural connectivity shapes cross-sectional grey matter differences across illness stages

We first evaluated the performance of the three CDMs in capturing cross-sectional differences in regional GMV. In all datasets, the CDM_SCw_ model yielded more accurate predictions of cross-sectional empirical GMV case-control differences (. 46 < *r* < .57; Fig2E-H) when compared with the CDM_SC_ and CDM_FCw_ models (*all r* < .35 ; Fig2E-H). For all data sets, the performance of the CDM_SCw_ was also significantly better than all three benchmark models (all *p* < 0.01). The CDM_SC_ and CDM_FCw_ generally did not surpass the performance of the benchmark models.

### Structural connectivity shapes longitudinal GMV changes

Having robustly demonstrated that cross-sectional grey matter differences at different illness stages are related to connectome structure, we next tested the implicit assumption of the CDM––that longitudinal GMV changes spread across axonal pathways––by considering longitudinal illness-related and medication-related changes in the FEP sample.

Predictions of the CDM_SCw_ model for illness-related grey matter changes at 3 and 12 months were correlated with empirical changes at *r* = .58 (Fig3E) and *r* = .52 (Fig3F), respectively, and both were significantly better than all three benchmark models (all *p* < .001). By comparison, correlations for the CDM_SC_ and CDM_FCw_ did not exceed *r* = .38 and only showed significantly better performance compared to the Null_smash_ and the Null_spin_ at 3 months, but not connectome benchmarks (all *p* > 0.05; Fig2E-F).

**Fig 3.**
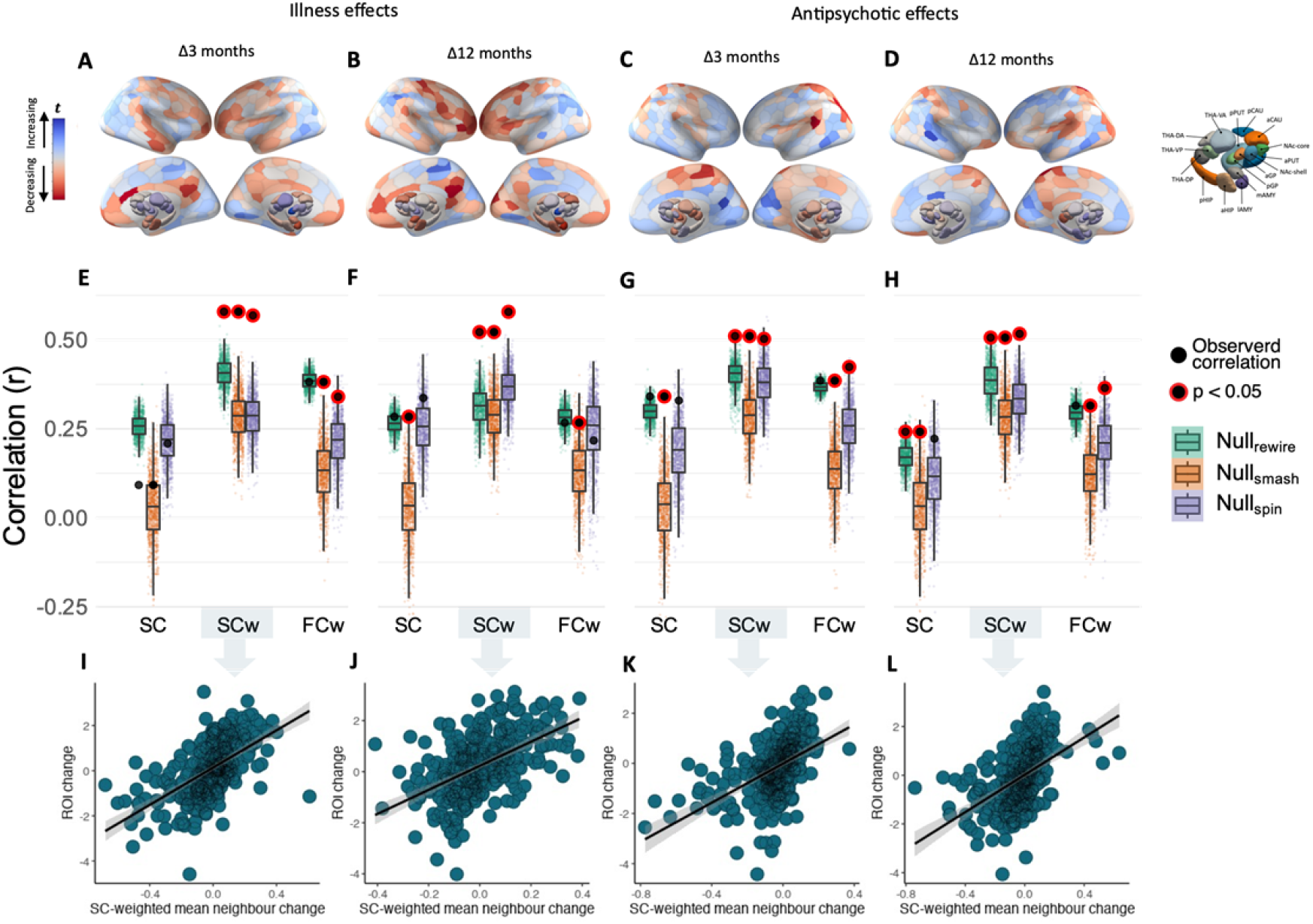
Longitudinal illness-related and antipsychotic-related GMV changes are constrained by connectome anatomy. **(A-D)** The contrast statistics for illness-related and antipsychotic-related contrasts mapped to a brain parcellation comprising 332 regions. **(E-H)** Performance of the equally weighted (CDM_SC_), structural connectivity-weighted (CDM_SCw_) and functional coupling-weighted (CDM_FCw_) models relative to the Null_smash_, Null_spin_, and Null_rewire_ benchmarks. Black circles indicate the observed rank correlations between predicted and actual regional deformation values for each model at each timepoint, with red borders indicating statistical significance. Note that the observed value used for comparison against the *Null*_*spin*_ models is different because the subcortex was excluded. **(I-L)** Scatterplots of the association between observed and predicted regional deformation values for the best performing CDM_SCw_ model at each timepoint.

Predictions of the CDM_SCw_ model for antipsychotic-related grey-matter changes were correlated with the empirical maps at *r* = .51 (Fig3E) for 3 months and *r* = .25 (Fig3F) for 12 months. This association was statistically significant when compared to all three null models (all *p* < 0.01; Fig3C-D). Associations at 3 months and 12 months were smaller for the CDM_SC_ (*r* = .34 and *r* = .24, respectively) and CDM_FCw_ models (*r* = .38 and *r* = .31, respectively). At 3 months, the CDM_SC_ and CDM_FCw_ models only showed significantly better performance than the Null_smash_ and Null_spin_ (*p* < .01) benchmarks. At 12 months, the CDM_SC_ model showed significantly better performance than the Null_smash_ and Null_rewire_ benchmarks and the CDM_SC_ model showed significantly better performance than the Null_smash_ and Null_spin_ benchmarks (*p* < .05). Thus, connectome structure represents a generic constraint on both illness-related and medication-related longitudinal GMV changes in psychosis.

### Epicentres of grey matter volume loss

We next used the NDM to simulate the dynamic spread of GMV loss from each individual brain region. Results using the Null_smash_ benchmark are presented below (Fig4C-J) and results using the Null_rewire_ benchmark are presented in the Supplement (FigS4). Across all cross-sectional comparisons, medial temporal lobe regions emerged as statistically significant epicentres (Fig4C-F). In particular, the anterior hippocampus was consistently implicated across all datasets (*p* < 0.05), surviving multiple comparison correction (*p*_FWE_ < 0.05) in the two schizophrenia samples. In the FEP dataset, additional epicentres were identified in bilateral occipital and temporal cortex, as well as hippocampus, amygdala, and posterior thalamic regions (Fig4C). In the EP dataset, additional epicentres were identified in temporal and posterior cingulate cortex, (Fig4D). In both schizophrenia samples, additional epicentres were identified in temporal cortex, amygdala, and posterior thalamic regions (Fig4E-F). Consistent results were obtained using the Null_rewire_ benchmark (FigS4).

**Fig 4.**
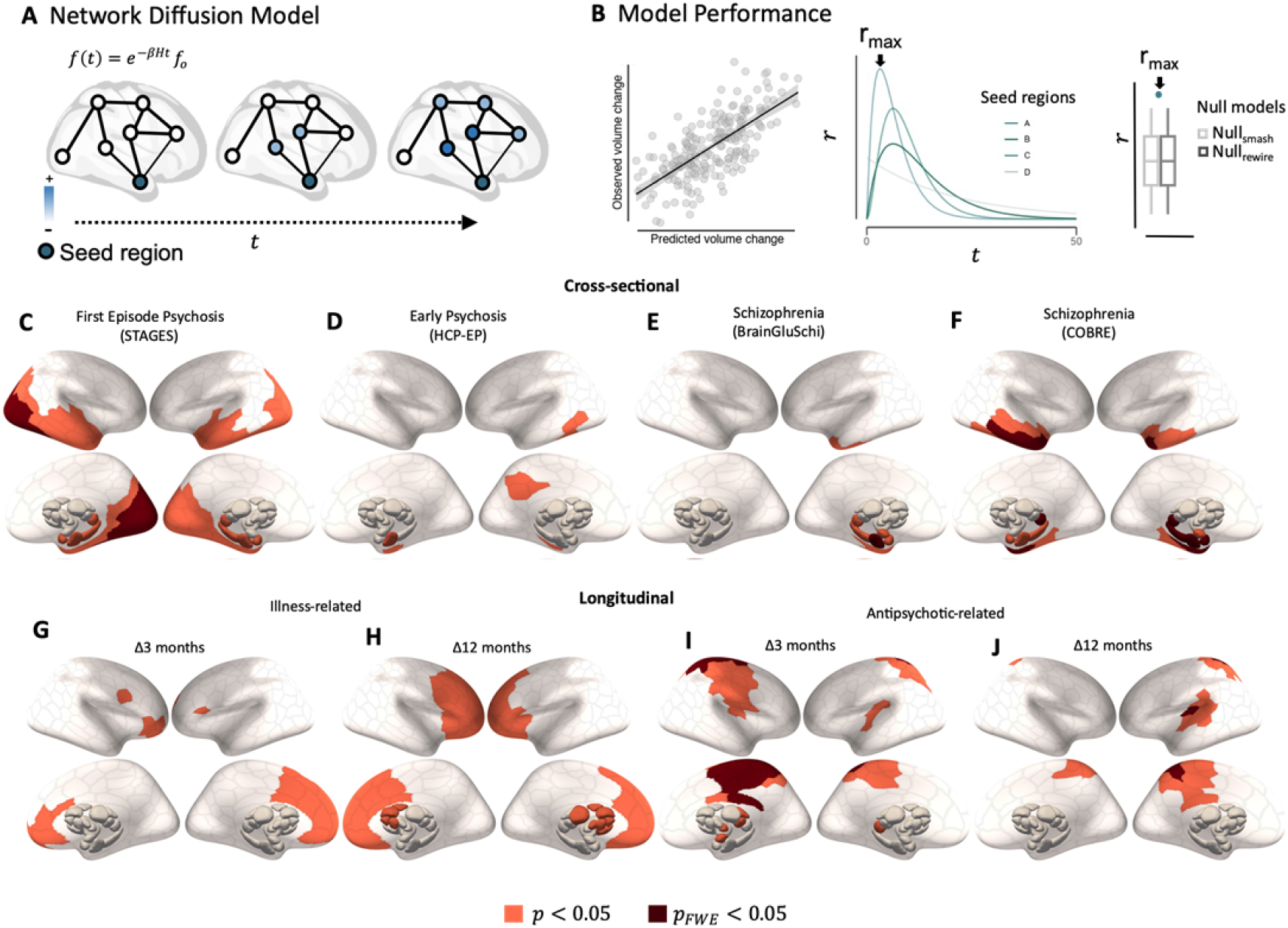
Regional epicentres of grey matter loss. **(A)** Epicentres were defined as potential sources of pathological volume loss from which GMV reductions spread (Blue) to affect structurally connected areas. To identify such regions, we simulated a spreading process using a Network Diffusion Model (NDM), **(B)** using each of the 332 parcellated regions as a seed, we retained the maximum correlation between the simulated and observed GMV abnormalities (*r*_*max*_). For each contrast, we then compared *r*_*max*_ values for each region to distribution of *r*_*max*_values from two benchmark null models accounting for spatial autocorrelations in the deformation maps (Null_smash_) and basic topological properties of the connectome (see Model evaluation (NDM)). Regional epicentres with significantly greater *r*_*max*_ than a spatially constrained null model (Orange = *p* < 0.05; Red = *p*_FWE_ < 0.05) are shown for cross-sectional **(C-F)** and longitudinal **(G-J)** effects. Results using Null_rewire_ benchmark models, and scatter plots of observed and predicted volume abnormalities are provided in Supplement (FigS3).

In the FEP sample, epicentres of longitudinal illness-related loss were identified in medial frontal regions at 3 months and progressed to include much of the frontal cortex, as well as striatal and thalamic regions, by 12 months (Fig4G-H). Comparison with the connectome-based null benchmarks were more conservative, but also implicated prefrontal regions (FigS4).

Epicentres of longitudinal antipsychotic-related GMV loss in FEP were identified in sensorimotor, cingulate and insula cortices, as well as thalamic and amygdala regions at 3 months, with the same cortical epicentres also identified at 12 months (Fig4I-J). These results were largely consistent when using the Null_rewire_ models (FigS4). Scatter plots of observed and predicted volume abnormalities for all contrasts are provided in the Supplement (FigS3).

### Robustness analyses

The magnitude and pattern of results remained consistent with our original findings after only considering individuals diagnosed with schizophrenia or schizophreniform disorder, indicating that diagnostic heterogeneity of the FEP and EP samples did not substantially impact our findings (FigS6).

To ensure that the wide-spread changes in white-matter integrity often reported in patients^50-53^ did not affect model estimates of the network-based spread of pathology, we replicated our findings using structural and functional connectomes derived from the FEP patient sample rather than the independent healthy control sample (FigS7). We also replicated the results using a representative structural connectome derived from the healthy control sample in the FEP study (FigS8).

Finally, our findings were consistent when using VBM instead of DBM (FigS9), and when applying global signal regression (GSR) on subject-level FC matrices before computing the group average FC matrix (FigS10).

## Discussion

The mechanisms driving spatially patterned grey matter volume (GMV) changes in psychotic illness have thus far been unclear. We have used a simple coordinated deformation model (CDM) to confirm that, across both early and later stages of illness, cross-sectional GMV changes are shaped by the topology and strength of structural, but not functional, connectivity between brain regions. We further found that both illness-related and antipsychotic-related longitudinal changes are constrained by structural connectivity, indicating that the temporal evolution of brain changes in the illness is also constrained by the brain’s axonal pathways. Moreover, using a network diffusion model (NDM) to simulate the spread of pathology from different brain regions, we identified the anterior hippocampus as a putative epicentre of volume loss across all illness stages and further showed a dynamic progression of epicentres of dynamic grey matter loss from posterior to anterior areas, suggesting that the pathological burden within temporal and prefrontal systems increases as the illness progresses.

### Structural connectivity constrains GMV changes in psychotic illness

The strength and topology of the structural connectome shaped the spatial pattern of volume abnormalities across both early and late stages of illness. Our findings in established schizophrenia align with previous research using the CDM to show that structural connectivity constrains the spatial patterning of cross-sectional GMV differences in people with established schizophrenia^31^. This earlier result was observed using the CDM_SC_ model considered here. In our analysis, we found that the strength of structural connectivity between regions modulates coupled GMV differences within structurally connected neighbourhoods, given that the CDM_SCw_ showed clearly superior performance to the CDM_SC_ and CDM_FCw_ models in all datasets. This result indicates that GMV differences are more tightly coupled between areas with high structural connectivity. Critically, our findings show that network constraints on cross-sectional GMV differences cannot be explained by antipsychotic medication, as our FEP sample was antipsychotic-naïve at the baseline scan. Moving beyond cross-sectional differences, our longitudinal analysis further demonstrates that both illness-related and antipsychotic-related changes in GMV are constrained by connectome architecture.

These results are in line with a spreading process in which pathology propagates across axonal connections. The precise mechanisms driving this process remain unclear. While there is limited evidence for visible deposits of aggregated pathological proteins in psychotic illness, more subtle changes in protein homeostasis^54^ may occur in subsets of patients and spread to synaptically connected distant brain regions^55^. Alternatively, and given the commonly reported finding of functional brain alterations in psychotic disorders, dysfunction of one region may trigger abnormal activity in connected sites which, over time, may trigger structural changes as a result of aberrant neurotransmission or a loss of trophic support^20^. This process may be exacerbated by a breakdown of white-matter fibre integrity, which may further disrupt the inter-regional transport of trophic factors. Accordingly, widespread but subtle alterations in white matter have been repeatedly demonstrated in psychosis populations^56^, are anticorrelated with cortical thickness^25^, and may predate the transition to psychosis in high-risk samples^50,51^. Although our analyses suggest that using a structural connectome derived from a patient sample did not change the overall pattern of our findings, further work may investigate how coordinated GMV changes interact with white-matter pathology in patients.

An alternative explanation for our findings is that regions sharing a strong anatomical connection have a more similar molecular and cytoarchitectonic profile, resulting in a shared vulnerability to illness- or treatment-related changes^57-60^. Future research should examine associations between the strength of structural connectivity and shared molecular features such as receptor profiles, gene expression, and synaptic density in patient populations.

### The medial temporal lobe as an epicentre of grey matter differences in psychosis

Our NDM analysis indicated that the medial temporal lobe, and the anterior hippocampus in particular, is a putative source of GMV loss in psychosis. The hippocampus has repeatedly been implicated in the pathogenesis of psychosis. It has been linked to early neurodevelopmental aberrations^61-63^ and often shows lower levels of mRNA and protein markers of synaptic and dendritic function post-mortem^64,65^. Recent *in vivo* PET imaging studies have also identified a loss of synaptic vesicle proteins^66,67^. Multiple animal models and human studies have indicated that a primary dysfunction occurring within the hippocampus^68,69^, such as a loss of pyramidal cell inhibition, results in downstream brain abnormalities including disinhibition of striatal dopamine release^70,71^ and aberrant corticostriatothalamic functioning^72^. Other evidence suggests that dysregulation of glutamate neurotransmission beginning in the CA1 region^73,74^ initiates the transition to psychotic illness and eventuates in an atrophic process in which neuropil and interneurons are reduced in other medial temporal and structurally connected regions.

### Regional epicentres of longitudinal grey matter change dynamically evolve with illness progression

While the hippocampus was robustly implicated as a putative epicentre for cross-sectional GMV differences at different illness stages, our analysis of longitudinal changes in the FEP group identified putative epicentres in striatal and prefrontal areas. This contrasts the largely posterior focus of cortical epicentres for baseline GMV differences in this group, suggesting that the most pronounced longitudinal GMV changes occurring early in the illness affect the prefrontal cortex, which aligns with the greater involvement of striatal and prefrontal areas at the 12 compared to 3-month follow-up. These findings also accord with longitudinal studies in early-onset schizophrenia demonstrating a dynamic wave of volume contraction progressing from posterior to anterior regions,^75,76^ and other evidence of pronounced prefrontal GMV reductions in the earliest stages of illness^8,77-83^, which may reflect an exaggeration of normal neurodevelopmental processes^84,85^. Notably, these regional epicentres of illness-related GMV loss were distinct from epicentres of antipsychotic-related GMV loss, which were identified in somatosensory, motor, and posterior cingulate regions.

### Limitations and conclusions

Our findings depend on group-level summary metrics of brain volume and may not be representative of volume changes at the individual patient level, which can show substantial heterogeneity^86-88^. Subsequent work could look at whether using individual-level measures of brain volume and connectivity can improve model predictions. Moreover, given the complexity and practical challenges of conducting a prospective triple-blind randomised control MRI study in antipsychotic-naïve patients, the sample size of the longitudinal FEP sample is small (see also^89,90^ for a discussion of the representativeness of this sample). Replication of our longitudinal analysis in larger samples is thus warranted.

In summary, we identify a robust and central role for axonal connectivity as a conduit for the spread of pathology across early and late stages of psychotic illness, mirroring findings reported in neurodegenerative conditions. Our findings also align with animal models to suggest that medial temporal regions may play a critical role in the origins of brain dysfunction and indicate that the structural connectome represents a fundamental constraint on brain changes in psychosis, regardless of whether they are caused by illness or medication.

## Data Availability

All code and group-level data used in the study are available online at https://github.com/sidchop/NetworkDeformationModels
Individual/patient level data may be made available upon reasonable request to the authors

## References

1 Gur, R. E., Turetsky, B. I., Bilker, W. B. & Gur, R. C. Reduced Gray Matter Volume in Schizophrenia. Archives of General Psychiatry 56, 905–911, doi:10.1001/archpsyc.56.10.905 (1999).

2 Haijma, S. V. et al. Brain Volumes in Schizophrenia: A Meta-Analysis in Over 18 000 Subjects. Schizophrenia Bulletin 39, 1129–1138, doi:10.1093/schbul/sbs118 (2013).

3 Steen, R. G., Mull, C., Mcclure, R., Hamer, R. M. & Lieberman, J. A. Brain volume in first-episode schizophrenia. British Journal of Psychiatry 188, 510–518, doi:10.1192/bjp.188.6.510 (2006).

4 van Erp, T. G. M. et al. Cortical Brain Abnormalities in 4474 Individuals With Schizophrenia and 5098 Control Subjects via the Enhancing Neuro Imaging Genetics Through Meta Analysis (ENIGMA) Consortium. Biological Psychiatry 84, 644–654, doi:10.1016/j.biopsych.2018.04.023 (2018).

5 van Erp, T. G. M. et al. Subcortical brain volume abnormalities in 2028 individuals with schizophrenia and 2540 healthy controls via the ENIGMA consortium. Molecular Psychiatry 21, 547–553, doi:10.1038/mp.2015.63 (2016).

6 Gupta, C. N. et al. Patterns of Gray Matter Abnormalities in Schizophrenia Based on an International Mega-analysis. Schizophrenia Bulletin 41, 1133–1142, doi:10.1093/schbul/sbu177 (2015).

7 Vieira, S. et al. Neuroanatomical abnormalities in first-episode psychosis across independent samples: a multi-centre mega-analysis. Psychological Medicine 51, 340–350, doi:10.1017/s0033291719003568 (2021).

8 Pantelis, C. et al. Neuroanatomical abnormalities before and after onset of psychosis: a cross-sectional and longitudinal MRI comparison. The Lancet 361, 281–288, doi:10.1016/S0140-6736(03)12323-9 (2003).

9 Vita, A., De Peri, L., Deste, G. & Sacchetti, E. Progressive loss of cortical gray matter in schizophrenia: a meta-analysis and meta-regression of longitudinal MRI studies. Translational Psychiatry 2, e190–e190, doi:10.1038/tp.2012.116 (2012).

10 Akudjedu, T. N. et al. Progression of neuroanatomical abnormalities after first-episode of psychosis: A 3-year longitudinal sMRI study. Journal of Psychiatric Research 130, 137–151, doi:10.1016/j.jpsychires.2020.07.034 (2020).

11 Olabi, B. et al. Are there progressive brain changes in schizophrenia? A meta-analysis of structural magnetic resonance imaging studies. Biological Psychiatry 70, 88–96, doi:10.1016/j.biopsych.2011.01.032 (2011).

12 Andreasen, N. C. et al. Progressive Brain Change in Schizophrenia: A Prospective Longitudinal Study of First-Episode Schizophrenia. Biological psychiatry 70, 672–679, doi:10.1016/j.biopsych.2011.05.017 (2011).

13 Chopra, S. et al. Differentiating the Effect of Medication and Illness on Brain Volume Reductions in First-Episode Psychosis: A Longitudinal, Randomized, Triple-blind, Placebo-controlled MRI study. medRxiv (2020).

14 Liloia, D. et al. Updating and characterizing neuroanatomical markers in high-risk subjects, recently diagnosed and chronic patients with schizophrenia: A revised coordinate-based meta-analysis. Neuroscience & Biobehavioral Reviews 123, 83–103 (2021).

15 Bora, E. et al. Neuroanatomical abnormalities in schizophrenia: A multimodal voxelwise meta-analysis and meta-regression analysis. Schizophrenia Research 127, 46–57, doi:10.1016/j.schres.2010.12.020 (2011).

16 Fornito, A., Yücel, M., Patti, J., Wood, S. J. & Pantelis, C. Mapping grey matter reductions in schizophrenia: An anatomical likelihood estimation analysis of voxel-based morphometry studies. Schizophrenia Research 108, 104–113, doi:10.1016/j.schres.2008.12.011 (2009).

17 Del Re, E. C. et al. Anterior-posterior axis of hippocampal subfields across psychoses: A B-SNIP study. Biomarkers in Neuropsychiatry 5, 100037, doi:https://doi.org/10.1016/j.bionps.2021.100037 (2021).

18 Sporns, O., Tononi, G. & Kötter, R. The Human Null_rewire_: A Structural Description of the Human Brain. PLOS Computational Biology 1, e42, doi:10.1371/journal.pcbi.0010042 (2005).

19 Raj, A. & Powell, F. Models of Network Spread and Network Degeneration in Brain Disorders. Biological Psychiatry: Cognitive Neuroscience and Neuroimaging 3, 788–797, doi:10.1016/j.bpsc.2018.07.012 (2018).

20 Fornito, A., Zalesky, A. & Breakspear, M. The connectomics of brain disorders. doi:10.1038/nrn3901 (2015).

21 Seeley, W. W., Crawford, R. K., Zhou, J., Miller, B. L. & Greicius, M. D. Neurodegenerative diseases target large-scale human brain networks. Neuron 62, 42–52 (2009).

22 Zhou, J., Gennatas, E. D., Kramer, J. H., Miller, B. L. & Seeley, W. W. Predicting regional neurodegeneration from the healthy brain functional connectome. Neuron 73, 1216–1227 (2012).

23 Raj, A., Kuceyeski, A. & Weiner, M. A network diffusion model of disease progression in dementia. Neuron 73, 1204–1215, doi:10.1016/j.neuron.2011.12.040 (2012).

24 Shafiei, G. et al. Network structure and transcriptomic vulnerability shape atrophy in frontotemporal dementia. Brain, doi:10.1093/brain/awac069 (2022).

25 Di Biase, M. A. et al. Linking Cortical and Connectional Pathology in Schizophrenia. Schizophr Bull 45, 911–923, doi:10.1093/schbul/sby121 (2019).

26 Wannan, C. M. J. et al. Evidence for Network-Based Cortical Thickness Reductions in Schizophrenia. American Journal of Psychiatry, appi.ajp.2019.18040380, doi:10.1176/appi.ajp.2019.18040380 (2019).

27 Cauda, F. et al. The morphometric co-atrophy networking of schizophrenia, autistic and obsessive spectrum disorders. Hum Brain Mapp 39, 1898–1928, doi:10.1002/hbm.23952 (2018).

28 Jiang, Y. et al. Antipsychotics effects on network-level reconfiguration of cortical morphometry in first-episode schizophrenia. medRxiv, 2021.2001.2017.21249965, doi:10.1101/2021.01.17.21249965 (2021).

29 Hettwer, M. et al. Coordinated cortical thickness alterations across six neurodevelopmental and psychiatric disorders. Nature Communications 13, 1–14 (2022).

30 Cauda, F. et al. Brain structural alterations are distributed following functional, anatomic and genetic connectivity. Brain 141, 3211–3232 (2018).

31 Shafiei, G. et al. Spatial patterning of tissue volume loss in schizophrenia reflects brain network architecture. Biological psychiatry 87, 727–735 (2020).

32 Chopra, S. et al. Functional connectivity in antipsychotic-treated and antipsychotic-naive patients with first-episode psychosis and low risk of self-harm or aggression: a secondary analysis of a randomized clinical trial. JAMA psychiatry 78, 994–1004 (2021).

33 Francey, S. M. et al. Psychosocial Intervention With or Without Antipsychotic Medication for First-Episode Psychosis: A Randomized Noninferiority Clinical Trial. Schizophrenia Bulletin Open 1, doi:10.1093/schizbullopen/sgaa015 (2020).

34 Lewandowski, K. E., Bouix, S., Ongur, D. & Shenton, M. E. Neuroprogression across the early course of psychosis. Journal of psychiatry and brain science 5 (2020).

35 Bustillo, J. R. et al. Glutamatergic and neuronal dysfunction in gray and white matter: a spectroscopic imaging study in a large schizophrenia sample. Schizophrenia bulletin 43, 611–619 (2017).

36 Çetin, M. S. et al. Thalamus and posterior temporal lobe show greater inter-network connectivity at rest and across sensory paradigms in schizophrenia. Neuroimage 97, 117–126, doi:10.1016/j.neuroimage.2014.04.009 (2014).

37 Esteban, O. et al. MRIQC: Advancing the automatic prediction of image quality in MRI from unseen sites. PLOS ONE 12, e0184661, doi:10.1371/journal.pone.0184661 (2017).

38 Gaser, C. & Dahnke, R. CAT - A Computational Anatomy Toolbox for the Analysis of Structural MRI Data. HBM, 336–348 (2016).

39 Ashburner, J. et al. SPM12 manual. Wellcome Trust Centre for Neuroimaging, London, UK 2464 (2014).

40 Schwarz, D. & Kašpárek, T. Comparison of Two Methods for Automatic Brain Morphometry Analysis. Radioengineering 20 (2011).

41 Guillaume, B., Hua, X., Thompson, P. M., Waldorp, L. & Nichols, T. E. Fast and accurate modelling of longitudinal and repeated measures neuroimaging data. NeuroImage 94, 287–302, doi:10.1016/j.neuroimage.2014.03.029 (2014).

42 Schaefer, A. et al. Local-Global Parcellation of the Human Cerebral Cortex from Intrinsic Functional Connectivity MRI. Cerebral Cortex (New York, N.Y.: 1991) 28, 3095–3114, doi:10.1093/cercor/bhx179 (2018).

43 Tian, Y., Margulies, D. S., Breakspear, M. & Zalesky, A. Topographic organization of the human subcortex unveiled with functional connectivity gradients. Nature Neuroscience 23, 1421–1432, doi:10.1038/s41593-020-00711-6 (2020).

44 Murphy, K. & Fox, M. D. Towards a consensus regarding global signal regression for resting state functional connectivity MRI. Neuroimage 154, 169–173, doi:10.1016/j.neuroimage.2016.11.052 (2017).

45 Aquino, K. M., Fulcher, B. D., Parkes, L., Sabaroedin, K. & Fornito, A. Identifying and removing widespread signal deflections from fMRI data: Rethinking the global signal regression problem. Neuroimage 212, 116614 (2020).

46 Betzel, R. F. & Bassett, D. S. Specificity and robustness of long-distance connections in weighted, interareal connectomes. Proceedings of the National Academy of Sciences, 201720186, doi:10.1073/PNAS.1720186115 (2018).

47 Raj, A., Kuceyeski, A. & Weiner, M. A network diffusion model of disease progression in dementia. Neuron 73, 1204–1215, doi:10.1016/j.neuron.2011.12.040 (2012).

48 Raj, A. et al. Network diffusion model of progression predicts longitudinal patterns of atrophy and metabolism in Alzheimer’s disease. Cell reports 10, 359–369 (2015).

49 Pandya, S. et al. Predictive model of spread of Parkinson’s pathology using network diffusion. NeuroImage 192, 178–194 (2019).

50 Carletti, F. et al. Alterations in White Matter Evident Before the Onset of Psychosis. Schizophrenia Bulletin 38, 1170–1179, doi:10.1093/schbul/sbs053 (2012).

51 Di Biase, M. A. et al. White matter changes in psychosis risk relate to development and are not impacted by the transition to psychosis. Molecular Psychiatry, doi:10.1038/s41380-021-01128-8 (2021).

52 Kraguljac, N. V. et al. White matter integrity, duration of untreated psychosis, and antipsychotic treatment response in medication-naïve first-episode psychosis patients. Molecular Psychiatry 26, 5347–5356, doi:10.1038/s41380-020-0765-x (2021).

53 Kelly, S. et al. Widespread white matter microstructural differences in schizophrenia across 4322 individuals: results from the ENIGMA Schizophrenia DTI Working Group. Molecular psychiatry 23, 1261–1269 (2018).

54 Bradshaw, N. J. & Korth, C. Protein misassembly and aggregation as potential convergence points for non-genetic causes of chronic mental illness. Molecular psychiatry 24, 936–951 (2019).

55 Guo, J. L. & Lee, V. M. Cell-to-cell transmission of pathogenic proteins in neurodegenerative diseases. Nature medicine 20, 130–138 (2014).

56 Yao, L. et al. White matter deficits in first episode schizophrenia: An activation likelihood estimation meta-analysis. Progress in Neuro-Psychopharmacology and Biological Psychiatry 45, 100–106, doi:https://doi.org/10.1016/j.pnpbp.2013.04.019 (2013).

57 Arnatkeviciute, A. et al. Genetic influences on hub connectivity of the human connectome. Nature Communications 12, 4237, doi:10.1038/s41467-021-24306-2 (2021).

58 Fornito, A., Arnatkevičiūtė, A.] & Fulcher, B. D. Bridging the Gap between Null_rewire_ and Transcriptome. Trends Cogn Sci 23, 34–50, doi:10.1016/j.tics.2018.10.005 (2019).

59 Hansen, J. Y. et al. Mapping neurotransmitter systems to the structural and functional organization of the human neocortex. bioRxiv, 2021.2010.2028.466336, doi:10.1101/2021.10.28.466336 (2022).

60 Hansen, J. Y. et al. Local molecular and global connectomic contributions to cross-disorder cortical abnormalities. Nature communications 13, 1–17 (2022).

61 Lipska, B. K. & Weinberger, D. R. A neurodevelopmental model of schizophrenia: neonatal disconnection of the hippocampus. Neurotoxicity research 4, 469–475 (2002).

62 Rosso, I. M. et al. Obstetric risk factors for early-onset schizophrenia in a Finnish birth cohort. American Journal of Psychiatry 157, 801–807 (2000).

63 Bearden, C. E., Meyer, S. E., Loewy, R. L., Niendam, T. A. & Cannon, T. D. The neurodevelopmental model of schizophrenia: Updated. Developmental Psychopathology: Volume Three: Risk, Disorder, and Adaptation, 542–569 (2015).

64 Osimo, E. F., Beck, K., Reis Marques, T. & Howes, O. D. Synaptic loss in schizophrenia: a meta-analysis and systematic review of synaptic protein and mRNA measures. Molecular Psychiatry 24, 549–561, doi:10.1038/s41380-018-0041-5 (2019).

65 Harrison, P. J. & Eastwood, S. L. Neuropathological studies of synaptic connectivity in the hippocampal formation in schizophrenia. Hippocampus 11, 508–519 (2001).

66 Radhakrishnan, R. et al. In vivo evidence of lower synaptic vesicle density in schizophrenia. Molecular Psychiatry, doi:10.1038/s41380-021-01184-0 (2021).

67 Onwordi, E. C. et al. Synaptic density marker SV2A is reduced in schizophrenia patients and unaffected by antipsychotics in rats. Nature Communications 11, 246, doi:10.1038/s41467-019-14122-0 (2020).

68 Weinberger, D. R. Implications of Normal Brain Development for the Pathogenesis of Schizophrenia. Archives of General Psychiatry 44, 660–669, doi:10.1001/archpsyc.1987.01800190080012 (1987).

69 Lodge, D. J. & Grace, A. A. Hippocampal dysregulation of dopamine system function and the pathophysiology of schizophrenia. Trends in pharmacological sciences 32, 507–513 (2011).

70 Grace, A. Dopamine system dysregulation by the hippocampus: implications for the pathophysiology and treatment of schizophrenia. Neuropharmacology 62, 1342–1348 (2012).

71 Modinos, G., Allen, P., Grace, A. A. & McGuire, P. Translating the MAM model of psychosis to humans. Trends Neurosci 38, 129–138, doi:10.1016/j.tins.2014.12.005 (2015).

72 Sabaroedin, K., Tiego, J. & Fornito, A. Circuit-based approaches to understanding corticostriatothalamic dysfunction across the psychosis continuum. Biological Psychiatry (2022).

73 Lieberman, J. et al. Hippocampal dysfunction in the pathophysiology of schizophrenia: a selective review and hypothesis for early detection and intervention. Molecular psychiatry 23, 1764–1772 (2018).

74 Schobel, S. A. et al. Imaging patients with psychosis and a mouse model establishes a spreading pattern of hippocampal dysfunction and implicates glutamate as a driver. Neuron 78, 81–93 (2013).

75 Vidal, C. N. et al. Dynamically Spreading Frontal and Cingulate Deficits Mapped in Adolescents With Schizophrenia. Archives of General Psychiatry 63, 25–34, doi:10.1001/archpsyc.63.1.25 (2006).

76 Thompson, P. M. et al. Mapping adolescent brain change reveals dynamic wave of accelerated gray matter loss in very early-onset schizophrenia. Proceedings of the National Academy of Sciences 98, 11650–11655, doi:10.1073/pnas.201243998 (2001).

77 Sun, D. et al. Progressive Brain Structural Changes Mapped as Psychosis Develops in ‘At Risk’ Individuals. Schizophrenia research 108, 85–92, doi:10.1016/j.schres.2008.11.026 (2009).

78 Sun, D. et al. Brain surface contraction mapped in first-episode schizophrenia: a longitudinal magnetic resonance imaging study. Molecular Psychiatry 14, 976–986, doi:10.1038/mp.2008.34 (2009).

79 Jalbrzikowski, M. Association of Structural Magnetic Resonance Imaging Measures With Psychosis Onset in Individuals at Clinical High Risk for Developing Psychosis: An ENIGMA Working Group Mega-analysis. JAMA Psychiatry 78, 753–766, doi:10.1001/jamapsychiatry.2021.0638 (2021).

80 Cannon, T. D. et al. Progressive reduction in cortical thickness as psychosis develops: a multisite longitudinal neuroimaging study of youth at elevated clinical risk. Biol Psychiatry 77, 147–157, doi:10.1016/j.biopsych.2014.05.023 (2015).

81 Chung, Y. et al. Cortical abnormalities in youth at clinical high-risk for psychosis: Findings from the NAPLS2 cohort. NeuroImage: Clinical 23, 101862, doi:https://doi.org/10.1016/j.nicl.2019.101862 (2019).

82 Del Re, E. C. et al. Baseline Cortical Thickness Reductions in Clinical High Risk for Psychosis: Brain Regions Associated with Conversion to Psychosis Versus Non-Conversion as Assessed at One-Year Follow-Up in the Shanghai-At-Risk-for-Psychosis (SHARP) Study. Schizophrenia Bulletin 47, 562–574 (2021).

83 Collins, M. A. et al. Accelerated cortical thinning precedes and predicts conversion to psychosis: The NAPLS3 longitudinal study of youth at clinical high-risk. Mol Psychiatry, doi:10.1038/s41380-022-01870-7 (2022).

84 Cropley, V. L. et al. Accelerated Gray and White Matter Deterioration With Age in Schizophrenia. Am J Psychiatry 174, 286–295, doi:10.1176/appi.ajp.2016.16050610 (2017).

85 Gogtay, N., Vyas, N. S., Testa, R., Wood, S. J. & Pantelis, C. Age of onset of schizophrenia: perspectives from structural neuroimaging studies. Schizophr Bull 37, 504–513, doi:10.1093/schbul/sbr030 (2011).

86 Wolfers, T. et al. Mapping the Heterogeneous Phenotype of Schizophrenia and Bipolar Disorder Using Normative Models. JAMA Psychiatry, doi:10.1001/jamapsychiatry.2018.2467 (2018).

87 Lv, J. et al. Individual deviations from normative models of brain structure in a large cross-sectional schizophrenia cohort. Molecular Psychiatry 26, 3512–3523, doi:10.1038/s41380-020-00882-5 (2021).

88 Segal, A. et al. Regional, circuit, and network heterogeneity of brain abnormalities in psychiatric disorders. medRxiv, 2022.2003.2007.22271986, doi:10.1101/2022.03.07.22271986 (2022).

89 Leucht, S. et al. Clinical implications of Brief Psychiatric Rating Scale scores. British Journal of Psychiatry 187, 366–371, doi:10.1192/bjp.187.4.366 (2005).

90 Henry, L. P. et al. The EPPIC Follow-Up Study of First-Episode Psychosis: Longer-Term Clinical and Functional Outcome 7 Years After Index Admission. The Journal of Clinical Psychiatry 71, 716–728, doi:10.4088/JCP.08m04846yel (2010).

