## Supplement for "Network-based spreading of grey matter changes across different stages of psychosis"

***1A. Description of each dataset used***

***1B. MRI acquisition parameters***

***1C. DBM processing***

***1D. DWI processing***

***1E. fMRI processing***

***1F. FDR-Corrected and uncorrected voxel-level DBM t-statistic maps for each contrast***

***1G. FDR-Corrected and uncorrected voxel-level VBM t-statistic maps for each contrast***

***1H. Benchmark null models for the Coordinated Deformation Model (CDM)***

***1I. Further information Network Diffusion Model (NDM) and benchmark null models***

***1J. Data-driven epicentre mapping methods***

***FigS2. Data-driven epicentre region identification using different null models.***

***FigS3. Scatter plots of observed and predicted GMV alterations using the best seed across the whole brain.***

***FigS4. NDM epicentres using*** $\mathrm{Null}_{\mathrm{rewire}}$ ***nulls***

***FigS5. NDM epicentres using*** $\mathrm{Null}_{\mathrm{rewire}}$ ***nulls which do not preserve distance rules.***

***FigS6. Replication of results after excluding patients with non-schizophrenia diagnosis.***

***FigS7. Replication of results using representative structural and functional connectomes from the First Episode Psychosis (STAGES) patient population.***

***FigS8. Replication of results using alterative representative structural and functional connectomes.***

***FigS9. Replication of results using Voxel Based Morphometry.***

***FigS10. Replication of results after scaling SC and FC weights.***

***FigS11. Replication of results applying Global Signal Regression to FC data.***

### 1A. Description of each dataset used

*STAGES (First Episode Psychosis; FEP)*

We recruited 62 people aged 15-25 years (46% female) who were experiencing FEP. All patients had minimal previous exposure to antipsychotic medication (<7 days of use or lifetime 1750 mg chlorpromazine equivalent exposure) and a duration of untreated psychosis of less than 6 months. At baseline, patients were randomised to one of two groups: one given antipsychotic medication (risperidone or paliperidone) plus intensive psychosocial therapy and the other given placebo plus intensive psychosocial therapy. For both groups, the treatment period spanned 6-months. MRI was conducted at baseline, 3 months, and 12 months post-intake. The randomisation phase of the study terminated at 6 months, so patients in either the antipsychotic or placebo group could have received antipsychotic medication and ongoing psychosocial interventions after 6 months. In practice, four patients in the placebo group commenced antipsychotic medication in this intervening period, in addition to the four patients who had commenced at the 3-month timepoint. Thus, between the 3-month and 12-month scan, a total of eight patients in the placebo group commenced antipsychotic medication and were removed from the analysis. All patients in the antipsychotic continued medication with varying degrees of exposure. To ensure that our results were not dependant on inclusion of patients without schizophrenia, we repeated the primary analysis after only including individuals diagnosed with schizophrenia or schizophreniform disorder (see *Robustness analyses*).

A matched healthy control group comprising 27 individuals with no history of psychiatric or neurological diagnosis was also recruited and scanned alongside the patient groups. Demographic details of this sample are provided in Table 1. Further sample characteristics and details about research and safety protocols can be found elsewhere[1,2]. Ethical approval for the study was granted by the Melbourne Health Human Research Ethics Committee (MHREC:2007.616).

*Human Connectome Project Early Psychosis (Early Psychosis; EP)*

This ongoing study is acquiring brain MRI in a cohort of people with a psychosis-spectrum disorder and within the first 3 years of the onset of psychotic symptoms. The dataset also includes healthy control participants and the data release used here (Release 1.1) comprises 140 patients and 63 controls. All subjects are scanned across three sites based in the USA. Detailed inclusion and exclusion criteria for the dataset is described elsewhere[3]. In the current study, we used a subset of 121 patients and 57 controls who passed quality control and had complete and useable data. To ensure that our results were not dependant on inclusion of patients without schizophrenia, we repeated the primary analysis after only including individuals diagnosed with schizophrenia or schizophreniform disorder (see *Robustness analyses*).

*BrainGluSchi (Schizophrenia; SCZ-BGS)*

This is a publicly available dataset of brain MRI in a sample of 86 patients diagnosed with schizophrenia and 89 matched healthy controls. Additionally, all patients who were being treated with antipsychotics had to have been clinically stable on the same medications for >4 weeks. All patients were recruited from the University of New Mexico (UNM) Hospitals and all subjects were scanned at a single site. Detailed inclusion and exclusion criteria for the dataset is described elsewhere [4]. In the current study, we used a subset of 70 patients and 62 controls who passed quality control and had complete and useable data.

*COBRE (Schizophrenia; SCZ-COBRE)*

This is a publicly available dataset of brain MRI in a sample of 99 patients diagnosed with schizophrenia and 99 matched healthy controls. Additionally, all patients had to demonstrate retrospective and prospective clinical stability during three consecutive weekly visits and during each imaging assessment. All patients were scanned at a single site. Detailed inclusion and exclusion criteria for the dataset is described elsewhere[5]. In the current study, we used a subset of 66 patients and 72 controls who passed quality control and had complete and useable data.

*Independent healthy control sample*

We recruited a total of 356 healthy participants as part of a study conducted at Monash University, Australia. The participants were selected from a large cohort of 439 people as those with high-quality functional and diffusion MRI scans available. For further details, see Sabaroedin, et al. [6]. The study was conducted in accordance with the Monash University Human Research Ethics Committee (MUHREC: 2012001562).

### 1B. MRI acquisition parameters

*STAGES (First Episode Psychosis; FEP)*

Structural T1-weighted (T1w; MPRAGE) scans were acquired using a 3-T Siemens Trio Tim scanner with a 32-channel head coil at the Royal Children's Hospital in Melbourne, Australia. Image acquisition parameters at each timepoint were as follow: 176 sagittal slices, with a 1mm$^3$ voxel size, bandwidth 236 Hz/pixel, field of view (FOV) =256×256, matrix = 256×256×176, repetition time (TR)= 2300ms, echo time (TE) = 2.98ms and a 9° flip angle.

*Human* Null$_{rewire}$ *Project Early Psychosis (Early Psychosis; EP)*

Structural T1w (MPRAGE) scans were acquired using 3-T Siemens MAGNETOM Prisma scanners across three sites: Brigham and Women's Hospital, McLean Hospital, and Indiana University in USA. Brigham and Women's Hospital and Indiana University used a 32-channel head coil. McLean Hospital used a 64-channel head & neck coil, with the neck channels turned off. Image acquisition parameters were as follow: 208 sagittal slices, with a 0.8mm$^3$ voxel size, bandwidth 220 Hz/pixel, FOV =256×256, matrix = 256×256×208, TR = 2400ms, TE=2.22ms and flip angle = 8°.

*BrainGluSchi (Schizophrenia; SCZ-BGS) & COBRE (Schizophrenia; SCZ-COBRE)*

Structural T1w multi-echo MPRAGE scans were acquired using a 3-T Siemens TrioTim scanner with a 12-channel heal coil at Our Mind Research Network in New Mexico, USA. Image acquisition parameters were as follow: 176 sagittal slices, with a 1mm$^3$ voxel size, bandwidth 650 Hz/pixel, FOV =256×256 , matrix = 256×256×176, TR = 2530ms, number of echo's = 5, TE=/[1.64, 3.5, 5.36, 7.22, 9.08] ms and flip angle = 7°. The final image used for analysis was computed as the root mean square of the 5 images corresponding to each of the echo's.

*Independent healthy control sample*

Structural, diffusion and functional MRI data were acquired using a Siemens Skyra 3T scanner with a 32-channel head coil at Monash Biomedical Imaging in Melbourne, Australia. T1w structural scans were acquired using: 1 mm$^3$ isotropic voxels, TR = 2300ms, TE = 2.07ms, TI = 900ms, and a FOV of 256 mm.

Diffusion data were acquired using an interleaved acquisition with the following parameters: 2.5 mm3 voxel size, TR = 8800ms, TE = 110ms, FOV 240 mm, 60 directions with b = 3000 s/mm2, and seven b = 0 s/mm2 vol. In addition, a single b = 0 s/mm$^2$ was obtained with reversed phase encoding direction for susceptibility field estimation.

Multiband T2*-weighted whole-brain echo-planar images were acquired with a total of 620 functional volumes with 42 slices each were acquired per participant using an interleaved acquisition with the following parameters: TR = 754ms, TE = 21 milliseconds, flip angle of 50°, multiband acceleration factor of 3, FOV = 190mm, slice thickness of 3mm, and 3mm isotropic voxels. Participants were instructed to lie still in the scanner with eyes closed while maintaining wakefulness.

**1C. DBM processing**

For each participant, all scans were put through a spatial adaptive non-local means denoising filter, followed by internal resampling, bias correction, affine registration and the standard *SPM12* '*unified segmentation*'. After these initial pre-processing steps, the T1w images from all available timepoints were rigidly realigned to correct for differences in head position within-subject, and a subject-specific mean image was calculated and used as a reference in a subsequent realignment of all T1w images across all timepoints. The mean images were then normalised using the Diffeomorphic Anatomical Registration using Exponentiated Lie algebra algorithm (DARTEL; [7]) . The resulting spatial normalisation parameters were then applied to the bias-corrected individual images for all available timepoints. These native space images were then again realigned to a DARTEL normalised template, resulting in a voxel-wise map of Jacobian determinants, where the intensity of each voxel quantifies the amount of expansion or contraction required for registration to the template and a 3-mm FWHM smoothing kernel was applied to the Jacobian maps.

**1D. DWI processing**

We first implemented the *tractoflow*[8] pipeline, where the DWI data are denoised using the *dwidenoise* tool from *MRtrix3* and then skull-stripped using *FSL bet*. A N4 bias corrections then applied using *ANTs*[9] and the image was cropped using *Dipy* [10]. The *dwinormalise* tool from *MRtrix3*[11] was used to normalise the mean values in each image to approximately 1000. Data were then resampled to 1 mm isotropic spatial resolution and the *Dipy TensorModel* was used to estimate the Diffusion Tensor Image[12] at every voxel, with a weighted least squares method. The *csdeconv* package from *Dipy* was used to compute fibre orientation distributions (FOD), which represents the estimated orientation distribution of fibre structure at each voxel [13,14]. Peaks representing main diffusion directions were extracted from local maxima of each FOD's angular distribution. This FOD field was later used for tractography and to estimate the structural connectivity.

The T1w data were processed using the same protocol as DWI data for denoising, N4 bias correction, resampling, brain extraction, and cropping. The T1w data were then registered to the b0 image using non-linear ANTs [9]. Segmentation of grey matter, white matter, subcortex and cerebrospinal fluid was performed using *CIVIT* [15], and the resulting tissue partial volume estimate maps were used to compute the inclusion and exclusion masks as well as a grey/white matter interface mask used for seeding[8].

Probabilistic tracking was preformed using a particle filtering tractography algorithm[16] implemented in *Dipy*. Similar to anatomically constrained tractography[17], particle filtering tractography takes advantage of previously computed tissue maps to define areas where streamline can traverse. We set the maximum streamline length to 400mm and generated 10,000,000 streamlines. Default parameters were used for other local tracking options (step size = 0.5; maximum angle between 2 steps: 20)[18]. To create a SC matrix, streamlines were assigned to each of the closest regions in the parcellation within a 2-mm radius of the streamline endpoints[11], yielding undirected 332 × 332 connectivity matrices for all subject.

Importantly, most tractography algorithms are prone to false positives and do not directly index the quantitative strength of connections between pairs of regions[19,20]. We therefore implemented a state-of-the-art optimisation procedure, Convex Optimization Modelling for Microstructure Informed Tractography (COMMIT2), which has shown to be superior to other methods on key benchmarks derived from fibre-tracking phantoms [21]. COMMIT2 uses a forward model to recover the connectome with the minimum number of bundles that best explains the local axon density estimated from the DWI signal[21]. In doing so, COMMIT2 filters and re-weights pair-wise connections strengths and provides more biologically accurate quantitative estimates of connectivity. After optimised SC matrices were generated for each subject, we created a single group-average matrix by retaining connections if they appeared in at least $\tau$ subjects, where $\tau$ is the consensus threshold that results in a binary density comparable to that of a typical subject [22], and which was set to 38.6% for this sample. This threshold is computed separately for inter-/intra-hemispheric connections. Retained connections are assigned the corresponding group-average SC weight, resulting in a weighted group-average SC matrix. Finally, the SC weights from the group-average matrix were z-scored.

**1E. fMRI processing**

First, the fMRI data for each subject were processed in *FSL FEAT* [23] following a standard pipeline, which included removal of the first four volumes, rigid-body head motion correction, 3mm spatial smoothing to improve signal-to-noise ratio, and high-pass temporal filter of 75s to remove slow drifts. Subsequently, spatial independent component analysis was performed using *FSL MELODIC*[24]. These components were used as inputs for *FSL FIX* [25,26] an ICA-based denoising approach that uses an automated classifier to identify noise components and remove them from the data. This approach has been shown to successfully correct for motion and physiological noise, in addition to artifacts associated with multiband acceleration[26]. The *FSL-FIX* classifier was trained using an independent cohort of 25 individuals (13 males; mean age = 25.56 years), acquired using identical scanner and acquisition protocol, in which each of over 2000 components were manually labelled as signal or noise. The accuracy of the classifier in identifying nuisance components was verified in a subset of 15 individuals from our sample, yielding an accuracy estimate of 97%.

The time courses of components labelled as noise were used as nuisance regressors, along with 24 head motion parameters (6 rigid-body parameters, their backwards derivatives, and squared values of the 12 regressors). Given ongoing controversy around the application of global signal regression [27], we evaluated how this step affected our findings (see *Robustness analyses*). Denoised functional data were spatially normalized to the International Consortium for Brain Mapping 152 template in Montreal Neurological Institute (MNI) space using *ANTs* [9],via a three step method: 1) registration of the mean realigned functional scan to the skull-stripped high resolution anatomical scan via rigid-body registration; 2) spatial normalization of the anatomical scan to the MNI template via a nonlinear registration; and 3) normalization of functional scan to the MNI template using a single transformation matrix that concatenates the transforms generated in steps 1 and 2. We then computed whole brain FC matrices for each subject using pair-wise Pearson correlations between the timeseries from each of the 332 regions and took a mean FC matrix across the sample.

**1F. FDR-Corrected and uncorrected voxel-level DBM *t*-statistic maps for each contrast**

*A) STAGES (first episode psychosis)*

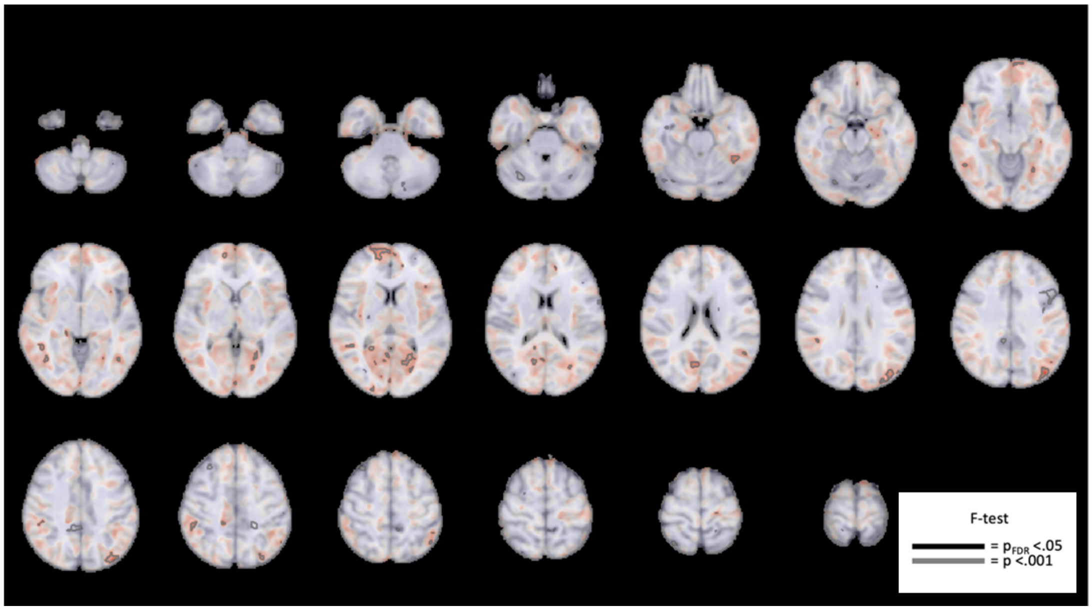


*B)* HCP-EP *(Early psychosis)*

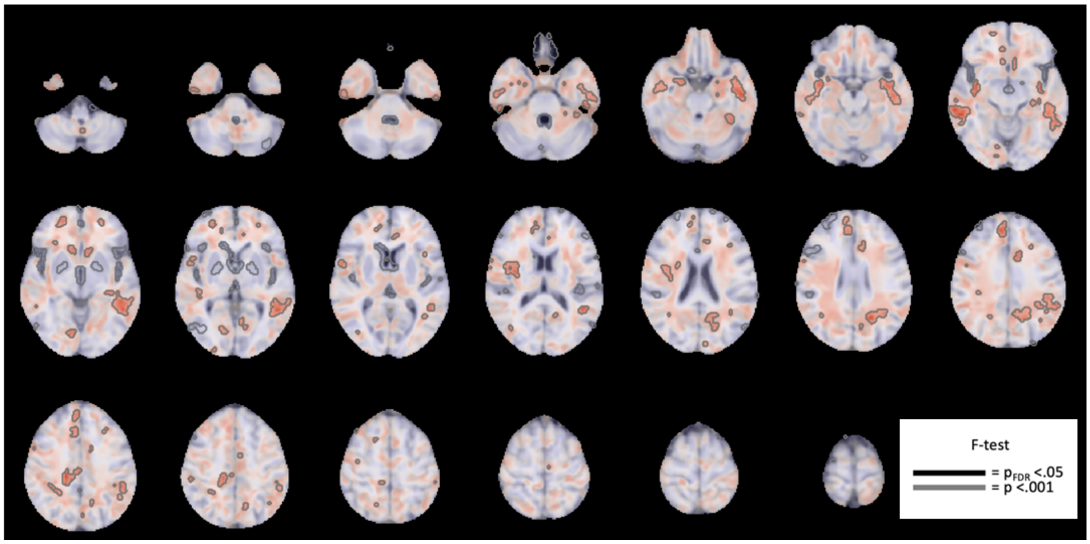

C) BrainGluSchi (schizophrenia)

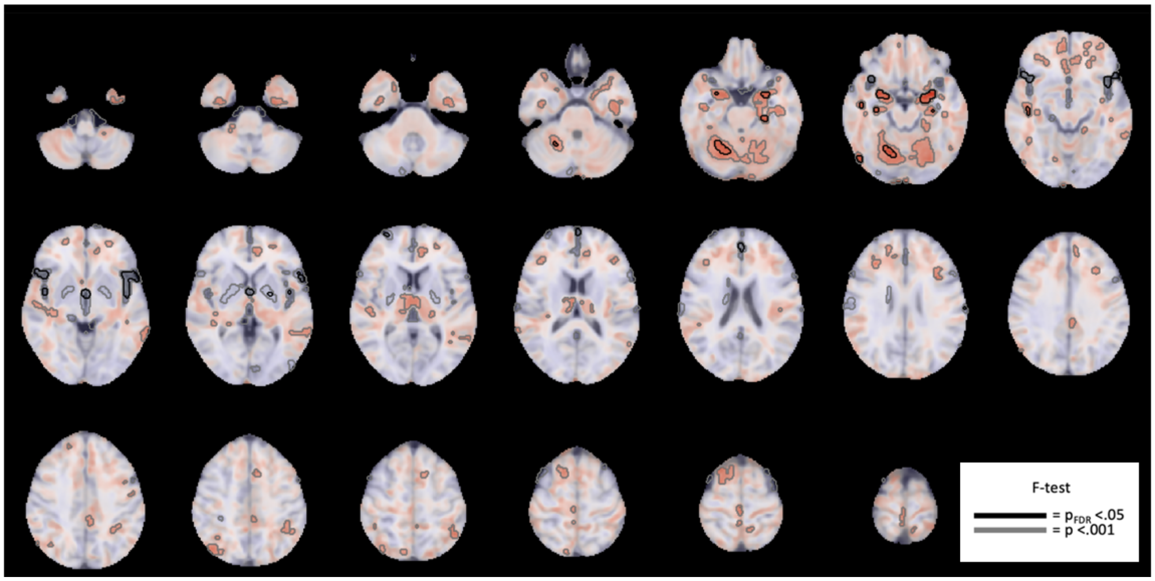


D) COBRE (schizophrenia)

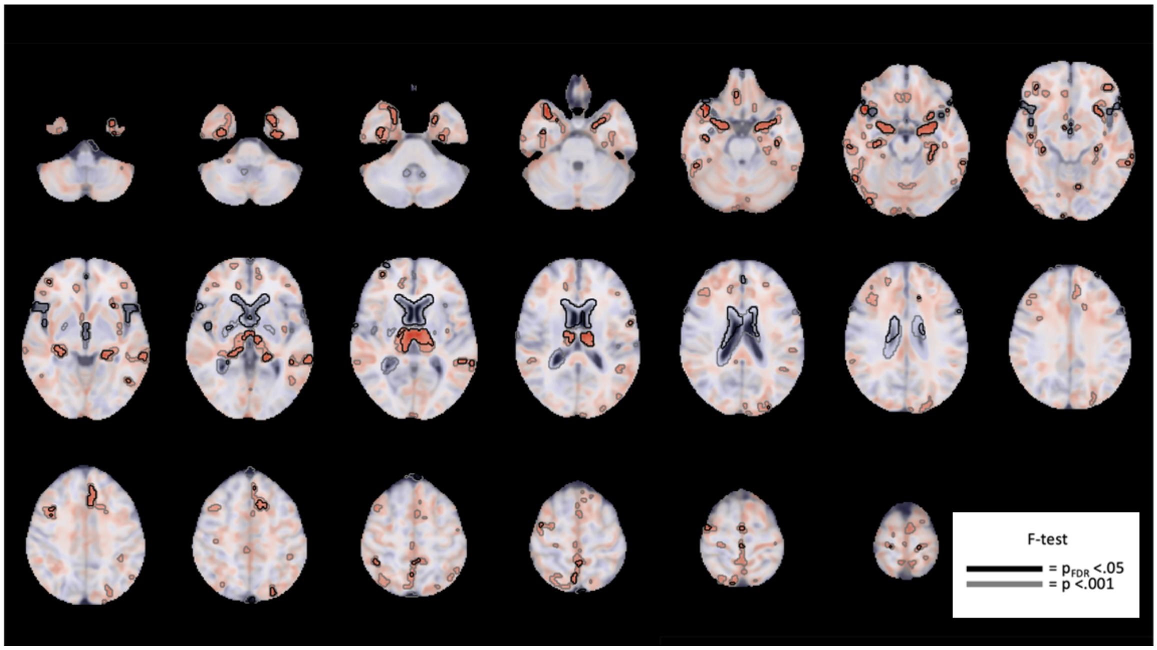

**1G. FDR-Corrected and uncorrected voxel-level VBM *t*-statistic maps for each contrast**

*A) STAGES (first episode psychosis)*

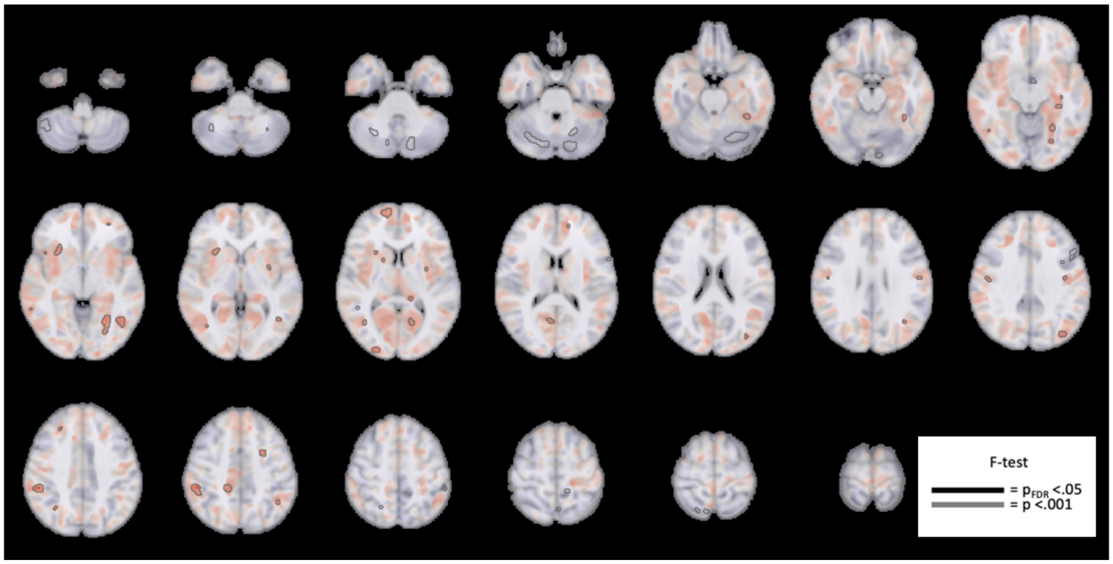


B) HCP-EP *(Early psychosis)*

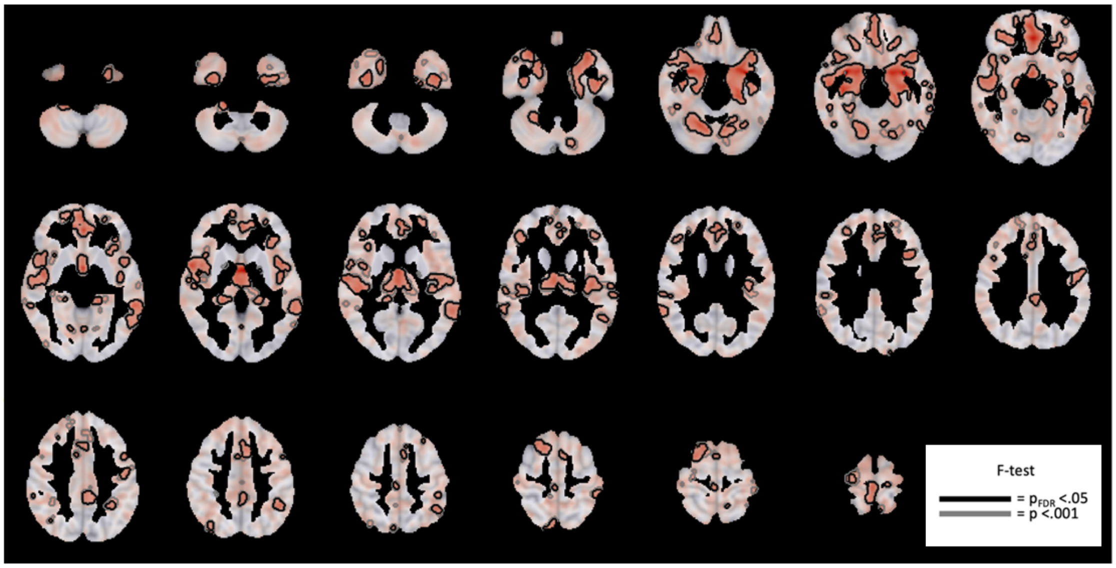

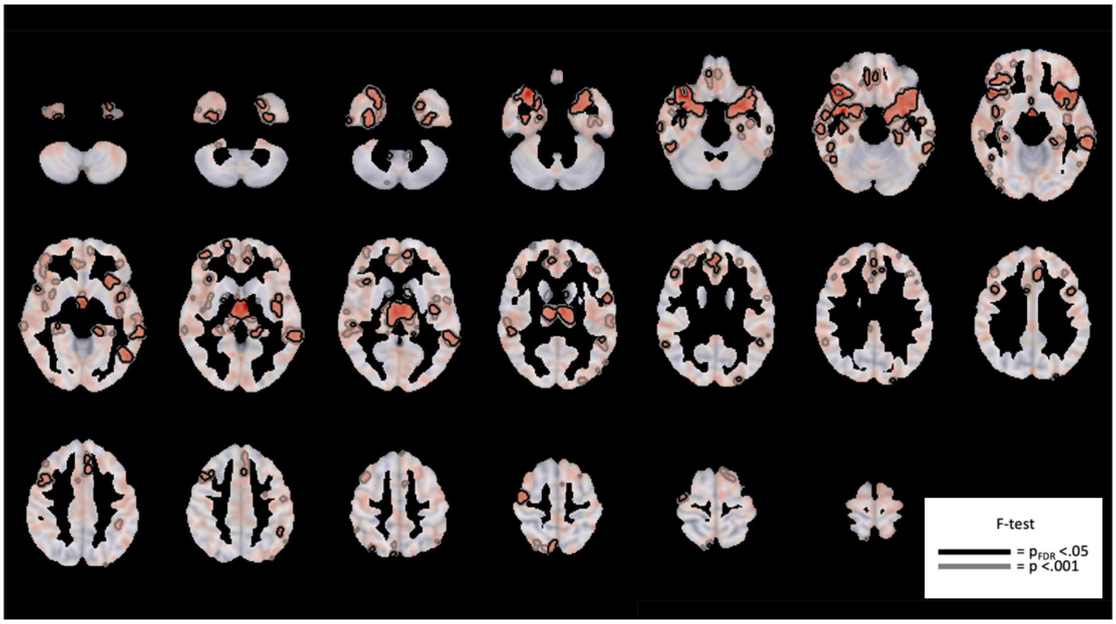


D) COBRE (schizophrenia)

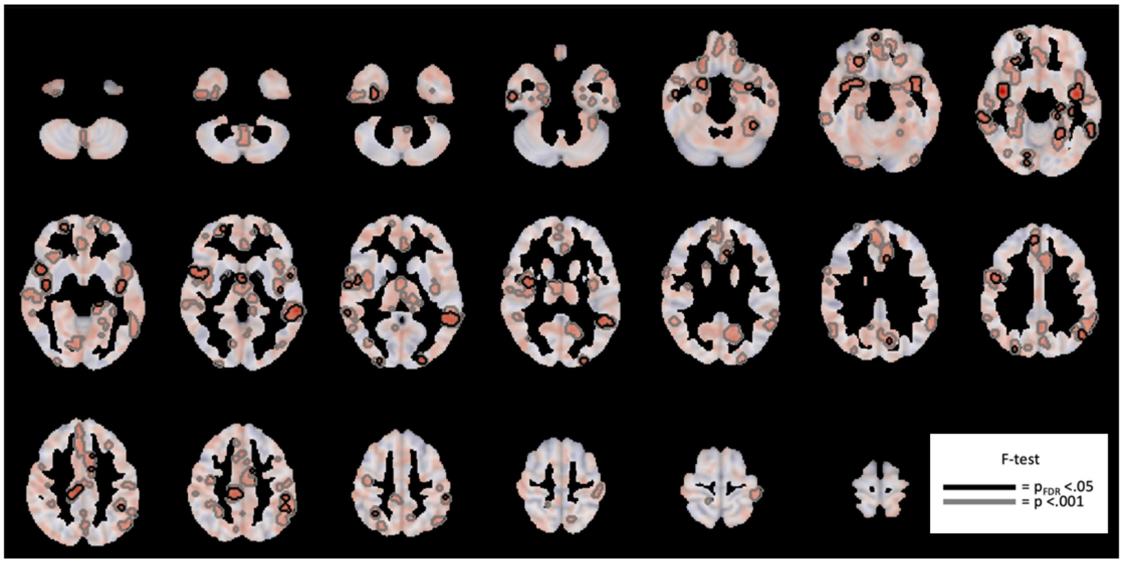

**1H. Benchmark null models for the Coordinated Deformation Model (CDM)**

Model performance was evaluated with respect to three null benchmark models. The first (Fig. 1D; $\text{Null}_{\text{smash}}$) and second (Fig. 1D; $\text{Null}_{\text{spin}}$) null models evaluated whether the observed findings were specific to the empirically observed pattern of grey matter deformations or were a generic property of the intrinsic spatial structure of the deformation maps. The two models differ in the way in which they account for the spatial structure present in the data. The first, $\text{Null}_{\text{smash}}$ approach used a parametric model to capture the spatial structure. Specifically, it relies on spatial variogram modelling to generate 1000 random spatial maps with a similar spatial autocorrelation to the observed deformation map, as implemented in the freely available toolbox *BrainSMASH*[28]; and with parameters (*ns=500*; *knn=2300*; *pv=70*) which resulted in null maps with variograms as close as possible to the empirical variogram across all contrasts. The second null benchmark, termed $\text{Null}_{\text{spin}}$, uses a spin-test to rotate region-level cortical $t$-values 1000 times[29]. The rotation was applied to one hemisphere and then mirrored for the other hemisphere. This benchmark is referred to as the $\text{Null}_{\text{spin}}$ null throughout the manuscript.

The primary advantage of the model-based method is that it can be applied to both cortical and subcortical data, however, it is not guaranteed to match the precise spatial autocorrelation of the empirical data. The spin test exactly preserves the empirical values and their spatial autocorrelation but is only applicable to cortex and also relies on certain approximations to account for the medial wall. In both cases, the 1000 surrogate values were used for inference on the observed performance metrics, with $p$-values quantified as the fraction of null values exceeding the observed correlation.

The third null model (Fig. 1D; $\text{Null}_{\text{rewire}}$) involved rewiring the structural connectome while preserving the degree sequence and length-weight relationship, and approximately preserving the edge-length distribution[30]. We used 10 distance bins and 50,000 edge swaps to generate 1000 rewired networks. These surrogate networks were used to test the hypothesis that any apparent network-based prediction of local grey matter change is specific to the actual topology of the connectome itself, and cannot be explained by basic network properties, such as regional variations in node degree or the spatial dependence of inter-regional connectivity. This benchmark null is referred to as the $\text{Null}_{\text{rewire}}$ null throughout the manuscript.

**1I. Further information Network Diffusion Model (NDM) and benchmark null models.**

The CDM model allowed us to determine whether brain connectivity shapes the spatial pattern of GMV alterations and whether SC or FC represents a stronger constraint on such patterns. However, it does not directly evaluate the mechanisms that link connectivity and GMV change, and it cannot identify whether individual regions act as sources or epicentres of volume loss. The NDM more directly tests the spreading hypotheses by simulating a passive diffusion process to model the spread of GMV alterations from specific seed regions. Thus, the NDM tests both the mechanism of spread, as well as the likely source, or epicentre, from which the spread may initiate. We therefore seeded the NDM using each of the 332 individual brain regions as a seed region for each seed region and each DBM contrast map (Fig1A), resulting in a total of 2656 simulations (332 regions × 8 contrasts). For each simulation, the measure of accuracy used to evaluate the NDM was the maximum correlation obtained across different time steps of the diffusion process ($r_{max}$; Fig4A) between the simulated and observed GMV loss. Note that, in principle, the NDM could be initialized using any combination of seed regions[31,32], but this can quickly result in a combinatorial explosion of alternative initial conditions for the model so we focused on testing specific hypotheses about individual brain regions as putative sources of GMV loss. Performance was evaluated separately for each hemisphere, but the family-wise error correction described below was evaluated across the whole brain and combined for both hemispheres when evaluating statistical significance of each brain region as an epicentre.

To evaluate the statistical significance of each region's $r_{max}$, we generated a null distribution of $r_{max}$ values using two benchmark models: the $\text{Null}_{\text{smash}}$ and the $\text{Null}_{\text{rewire}}$ null (see supplement section 1H for more information). To generate null $r_{max}$ values for each contrast using the $\text{Null}_{\text{smash}}$, we initiated the NDM from each brain region 1000 times and evaluated the correlation between the simulated GMV loss and the spatially constrained null GMV loss generated by the $\text{Null}_{\text{smash}}$. The $r_{max}$ from each iteration was retained, resulting in 1000 null $r_{max}$ values at each brain region. For each contrast and each brain region, the p-value was considered as the percentage of nulls $r_{max}$ null values greater than the observed $r_{max}$. To implement family-wise error (FWE) correction, for each contrast, the maximum brain-wide null $r_{max}$ from each of the 1000 iterations was used to construct a FWE-corrected null distribution[33]. The FWE-corrected p-value was considered as the percentage of FWE-corrected null $r_{max}$ values greater than the observed $r_{max}$. To evaluate significance using the $Null_{rewire}$ null model, we followed the same procedure describe above, but instead of varying the GMV volume loss, we varied the structural connectome at each of the 1000 iterations, using null connectomes generated using a rewiring method (Supplement section 1.8).

For most contrasts, the $Null_{smash}$ and $Null_{rewire}$ nulls identified consistent epicentres, although the connectome-based null benchmarks were more conservative in the analysis of longitudinal GMV change in the FEP sample, revealing a more circumscribed set of prefrontal regions compared to the $Null_{smash}$ benchmark. To understand the reasons for this discrepancy, we re-ran the epicentre analyses using a variant of the $Null_{rewire}$ null model that did not preserve the distance dependence of connectivity (FigS5), but still maintained other topological properties such as the node degree and edge-weight distributions[34]. The results were consistent with those obtained using the $Null_{smash}$ null (Fig4G-H) and implicated widespread frontal regions as epicentres of longitudinal GMV change. Thus, because these additional prefrontal regions emerge only when connection topology, but not the spatial dependence of connectivity, is preserved, their role as epicentres of longitudinal GMV change is largely due by their spatial proximity to the epicentres identified in the $Null_{rewire}$ analysis (Fig S4) rather than their profile of inter-regional connectivity.

*1J. Data-driven epicentre mapping methods*

As per Shafiei, et al. [35], we also implemented a data driven epicentre approach which defined such epicentres as areas showing high deformation that were also connected to regions showing high deformation. To identify such regions, for each region and each contrast, we rank-transformed and then took the mean of two values: (1) that region's extent of deformation; and (2) the mean of that region's neighbours' deformation, weighted by SC as in the $CDM_{SCw}$ model, given the superior performance of this model (see *Results*). Higher positive values on the resulting epicentre rank represent regions with high volume loss that are also connected to regions with high volume less. We then obtained a null ensemble of 1000 regional epicentre scores by repeating the same procedure after either rotating the regional deformation maps relative to the structural connectome to generate a distribution of null ranks at each region ($Null_{spin}$ benchmark). These 1000 null values were then used to quantify statistical significance of each region's epicentre score as the fraction of null values exceeding the observed rank score for a given regions (FigS2) [33]. We also characterised data driven epicentres using $Null_{smash}$ and $Null_{rewire}$ nulls (FigS2).

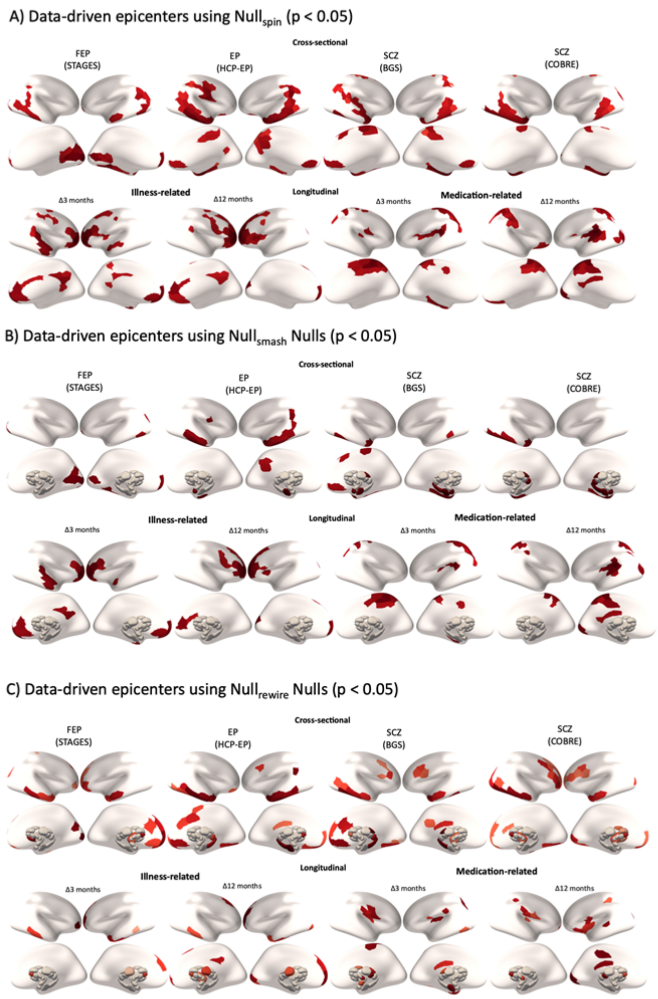


***FigS2 – Data-driven epicentre region identification using different null models.***

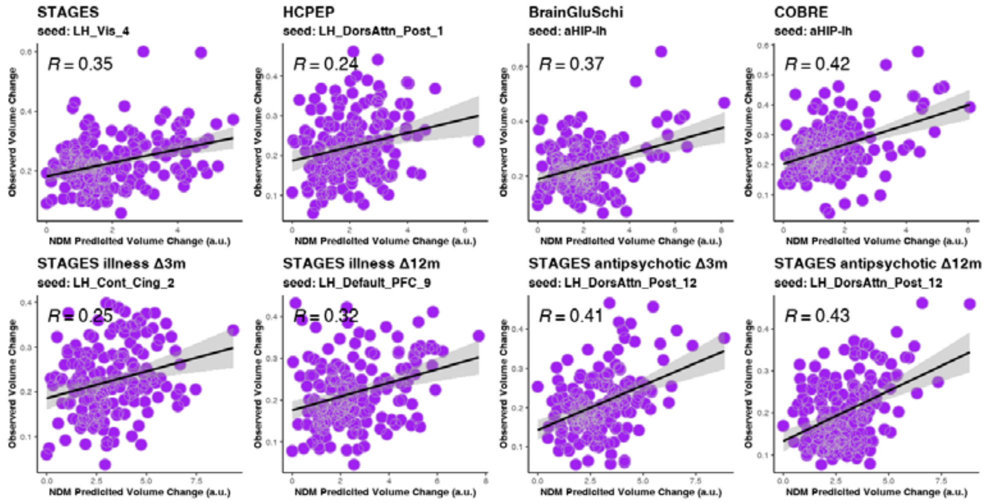


***FigS3 – Scatter plots of observed and predicted GMV alterations using the highest scoring seed across the whole brain.***

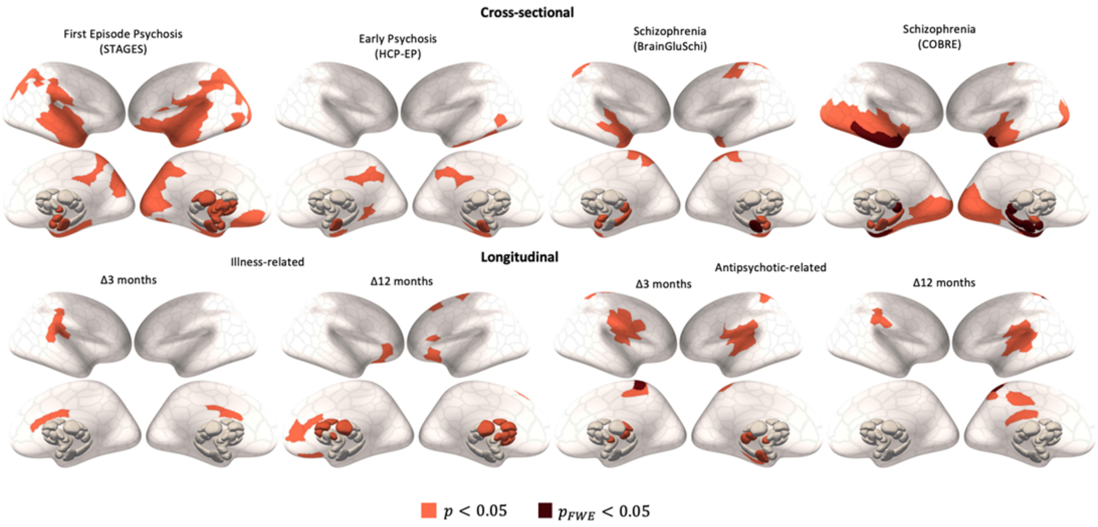


***FigS4 – NDM epicentres using Null$_{rewire}$ benchmark.***

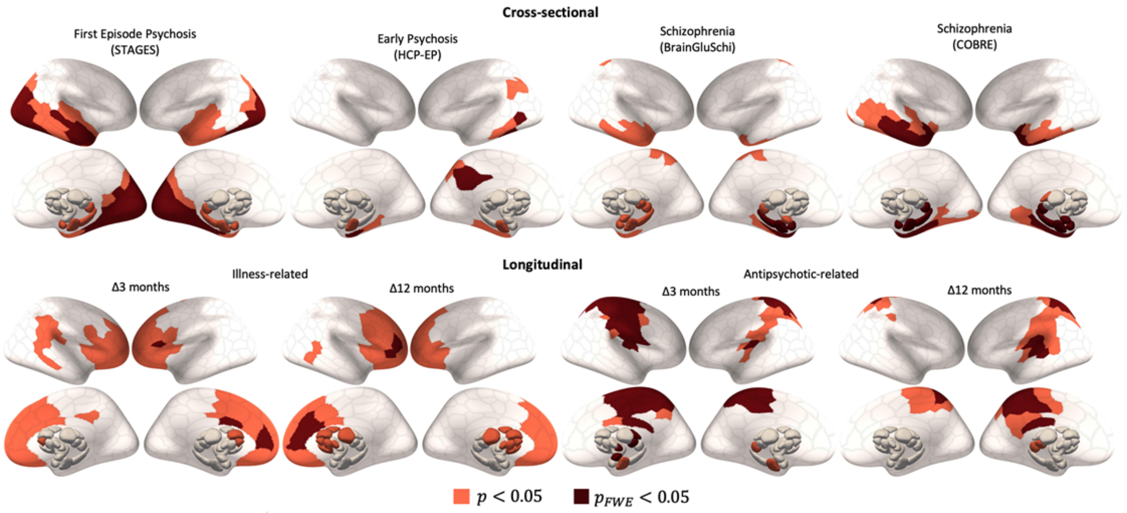


***FigS5 – NDM epicentres using Null$_{rewire}$ models which do not preserve distance rules.***

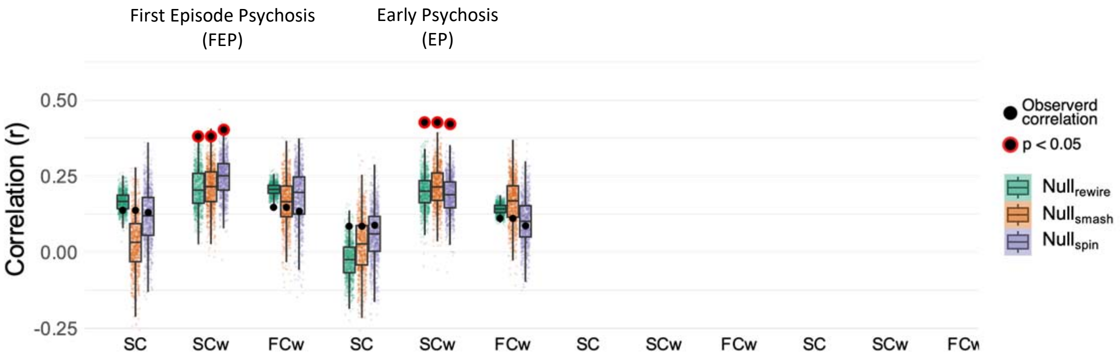


***FigS6 – Replication of cross-sectional results after excluding patients with non-schizophrenia diagnosis in FEP (STAGES) and EP (HCP-EP) samples.***

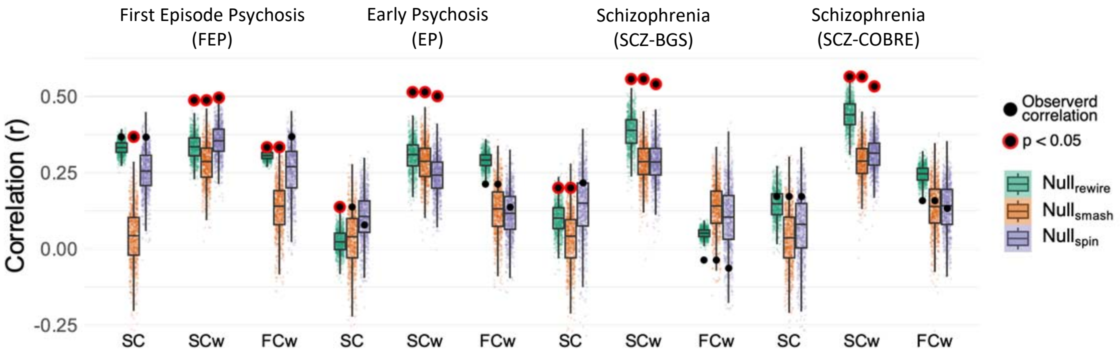


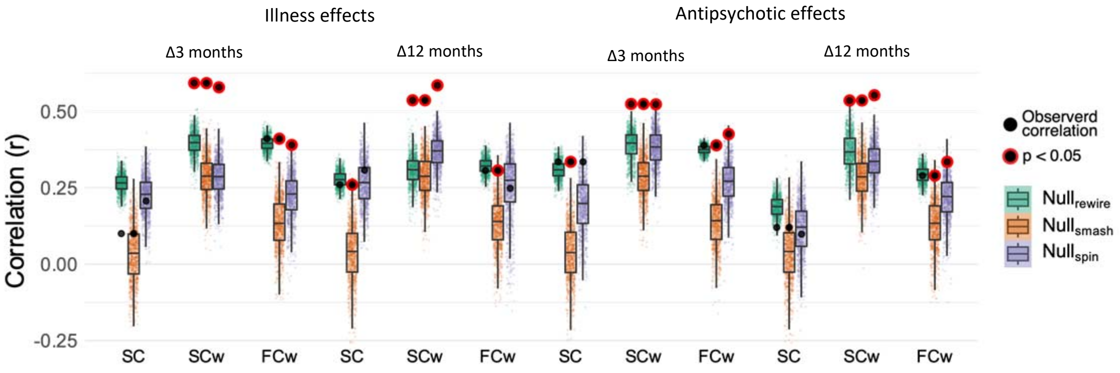


***FigS7 – Replication of results using representative structural and functional connectomes from the First Episode Psychosis (STAGES) patient population.***

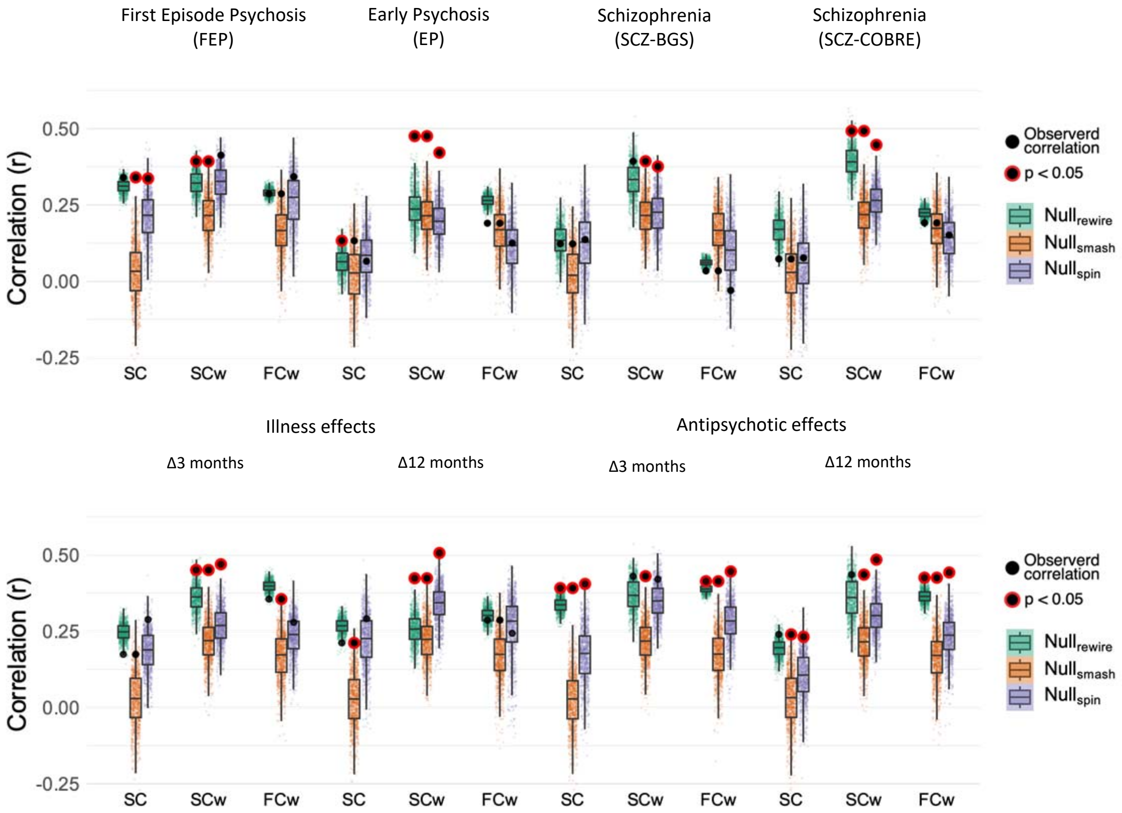


***FigS8 – Replication of results using alterative representative healthy structural and functional connectomes.***

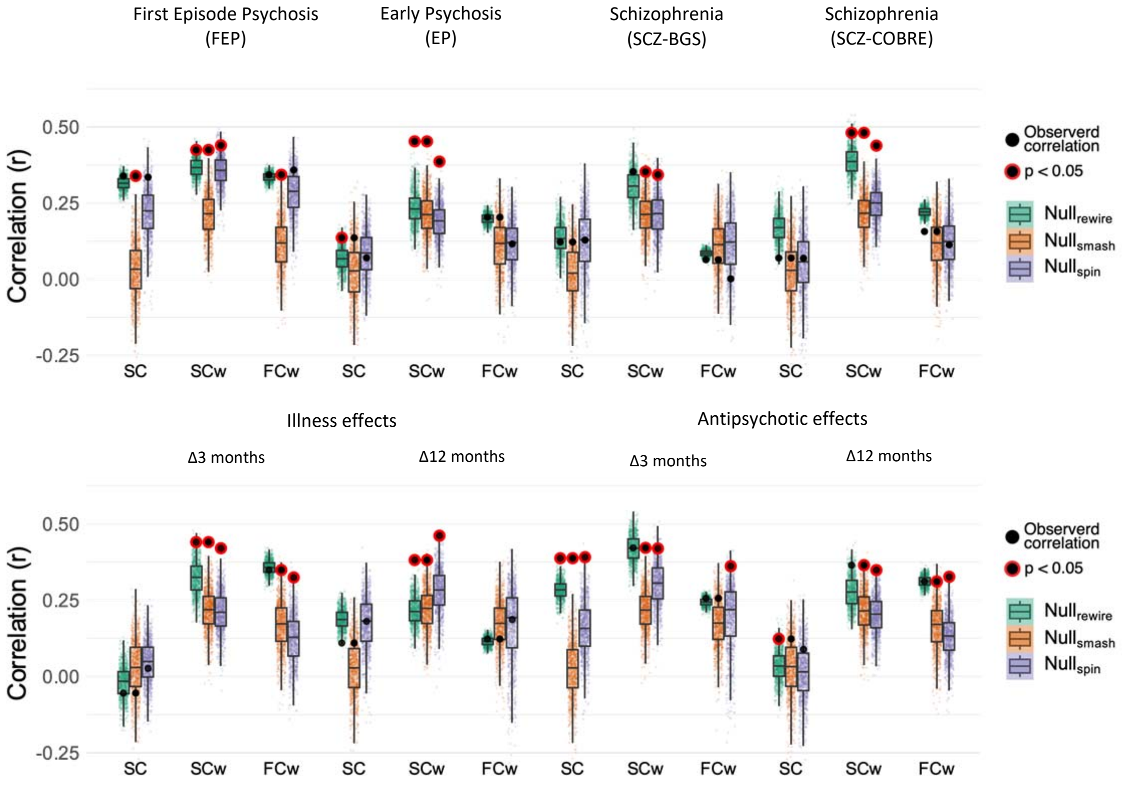


***FigS9 – Replication of results using Voxel Based Morphometry.***

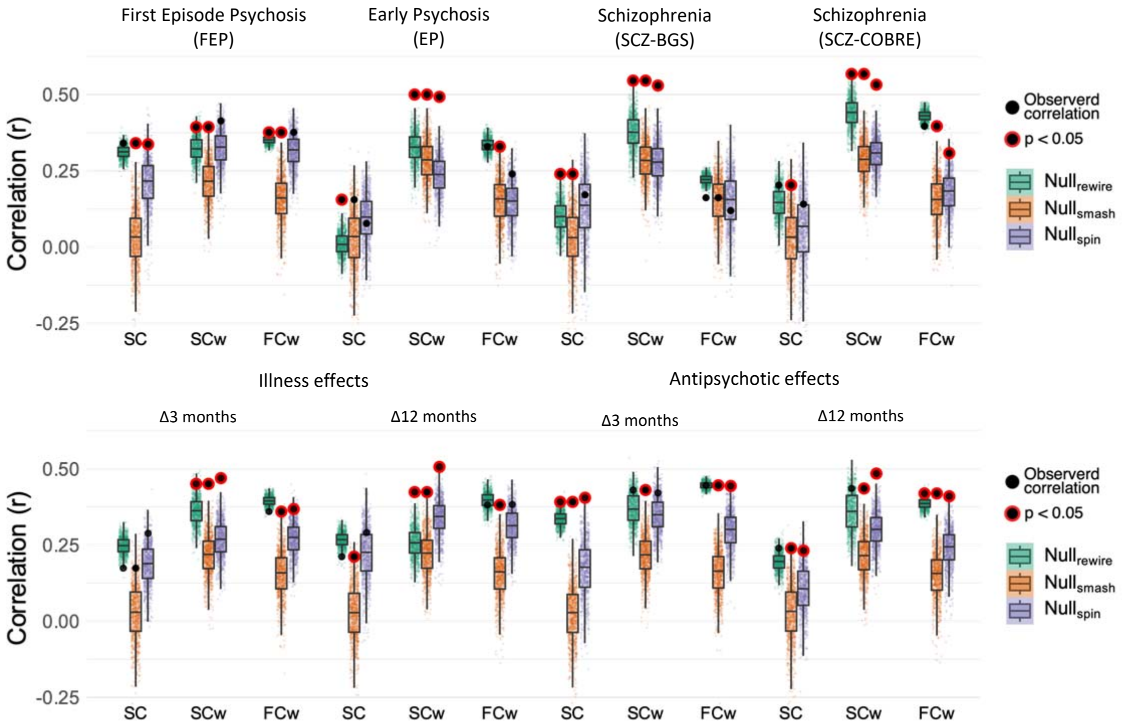


***FigS10 – Replication of results applying Global Signal Regression to FC data.***

**References**


1 Francey, S. M. *et al.* Psychosocial Intervention With or Without Antipsychotic Medication for First-Episode Psychosis: A Randomized Noninferiority Clinical Trial. *Schizophrenia Bulletin Open* **1**, doi:10.1093/schizbullopen/sgaa015 (2020).

2 O'Donoghue, B. *et al.* Staged treatment and acceptability guidelines in early psychosis study (STAGES): A randomized placebo controlled trial of intensive psychosocial treatment plus or minus antipsychotic medication for first-episode psychosis with low-risk of self-harm or aggression. Study protocol and baseline characteristics of participants. *Early Intervention in Psychiatry* **0**, doi:10.1111/eip.12716 (2019).

3 Lewandowski, K. E., Bouix, S., Ongur, D. & Shenton, M. E. Neuroprogression across the early course of psychosis. *Journal of psychiatry and brain science* **5** (2020).

4 Bustillo, J. R. *et al.* Glutamatergic and neuronal dysfunction in gray and white matter: a spectroscopic imaging study in a large schizophrenia sample. *Schizophrenia bulletin* **43**, 611-619 (2017).

5 Çetin, M. S. *et al.* Thalamus and posterior temporal lobe show greater inter-network connectivity at rest and across sensory paradigms in schizophrenia. *Neuroimage* **97**, 117-126, doi:10.1016/j.neuroimage.2014.04.009 (2014).

6 Sabaroedin, K. *et al.* Functional Connectivity of Corticostriatal Circuitry and Psychosis-like Experiences in the General Community. *Biological Psychiatry* **86**, 16-24, doi:10.1016/j.biopsych.2019.02.013 (2019).

7 Ashburner, J. A fast diffeomorphic image registration algorithm. *NeuroImage* **38**, 95-113, doi:10.1016/j.neuroimage.2007.07.007 (2007).

8 Theaud, G. *et al.* TractoFlow: A robust, efficient and reproducible diffusion MRI pipeline leveraging Nextflow & Singularity. *NeuroImage* **218**, 116889, doi:https://doi.org/10.1016/j.neuroimage.2020.116889 (2020).

9 Avants, B. B. *et al.* A reproducible evaluation of ANTs similarity metric performance in brain image registration. *Neuroimage* **54**, 2033-2044, doi:10.1016/j.neuroimage.2010.09.025 (2011).

10 Garyfallidis, E. *et al.* Dipy, a library for the analysis of diffusion MRI data. *Frontiers in Neuroinformatics* **8**, doi:10.3389/fninf.2014.00008 (2014).

11 Tournier, J. D. *et al.* MRtrix3: A fast, flexible and open software framework for medical image processing and visualisation. *NeuroImage* **202**, 116137, doi:https://doi.org/10.1016/j.neuroimage.2019.116137 (2019).

12 Basser, P. J., Mattiello, J. & LeBihan, D. MR diffusion tensor spectroscopy and imaging. *Biophys J* **66**, 259-267, doi:10.1016/s0006-3495(94)80775-1 (1994).

13 Tournier, J. D., Calamante, F. & Connelly, A. Robust determination of the fibre orientation distribution in diffusion MRI: Non-negativity constrained super-resolved spherical deconvolution. *NeuroImage* **35**, 1459-1472, doi:https://doi.org/10.1016/j.neuroimage.2007.02.016 (2007).

14 Descoteaux, M., Deriche, R., Knösche, T. R. & Anwander, A. Deterministic and probabilistic tractography based on complex fibre orientation distributions. *IEEE Trans Med Imaging* **28**, 269-286, doi:10.1109/tmi.2008.2004424 (2009).

15 Ad-Dab'bagh, Y. *et al.* in *Proceedings of the 12th annual meeting of the organization for human brain mapping.* (Florence, Italy).

16 Girard, G., Whittingstall, K., Deriche, R. & Descoteaux, M. Towards quantitative connectivity analysis: reducing tractography biases. *Neuroimage* **98**, 266-278, doi:10.1016/j.neuroimage.2014.04.074 (2014).

17 Smith, R. E., Tournier, J. D., Calamante, F. & Connelly, A. Anatomically-constrained tractography: improved diffusion MRI streamlines tractography through effective use

of anatomical information. *Neuroimage* **62**, 1924-1938, doi:10.1016/j.neuroimage.2012.06.005 (2012).

18 Al-Sharif, N. B., St-Onge, E., Theaud, G., Evans, A. C. & Descoteaux, M. Processing the diffusion-weighted magnetic resonance imaging of the PING dataset. *bioRxiv*, 2020.2011.2024.396549, doi:10.1101/2020.11.24.396549 (2020).

19 Maier-Hein, K. H. *et al.* The challenge of mapping the human connectome based on diffusion tractography. *Nature Communications* **8**, 1349, doi:10.1038/s41467-017-01285-x (2017).

20 Schilling, K. G. *et al.* Limits to anatomical accuracy of diffusion tractography using modern approaches. *Neuroimage* **185**, 1-11, doi:10.1016/j.neuroimage.2018.10.029 (2019).

21 Schiavi, S. *et al.* A new method for accurate in vivo mapping of human brain connections using microstructural and anatomical information. *Science advances* **6**, eaba8245-eaba8245, doi:10.1126/sciadv.aba8245 (2020).

22 Betzel, R. F., Griffa, A., Hagmann, P. & Mišić, B. Distance-dependent consensus thresholds for generating group-representative structural brain networks. *Netw Neurosci* **3**, 475-496, doi:10.1162/netn_a_00075 (2019).

23 Woolrich, M. W., Ripley, B. D., Brady, M. & Smith, S. M. Temporal autocorrelation in univariate linear modeling of FMRI data. *Neuroimage* **14**, 1370-1386, doi:10.1006/nimg.2001.0931 (2001).

24 Jenkinson, M., Beckmann, C. F., Behrens, T. E., Woolrich, M. W. & Smith, S. M. FSL. *Neuroimage* **62**, 782-790, doi:10.1016/j.neuroimage.2011.09.015 (2012).

25 Griffanti, L. *et al.* ICA-based artefact removal and accelerated fMRI acquisition for improved resting state network imaging. *Neuroimage* **95**, 232-247, doi:10.1016/j.neuroimage.2014.03.034 (2014).

26 Salimi-Khorshidi, G. *et al.* Automatic denoising of functional MRI data: combining independent component analysis and hierarchical fusion of classifiers. *Neuroimage* **90**, 449-468, doi:10.1016/j.neuroimage.2013.11.046 (2014).

27 Murphy, K. & Fox, M. D. Towards a consensus regarding global signal regression for resting state functional connectivity MRI. *Neuroimage* **154**, 169-173, doi:10.1016/j.neuroimage.2016.11.052 (2017).

28 Burt, J. B., Helmer, M., Shinn, M., Anticevic, A. & Murray, J. D. Generative modeling of brain maps with spatial autocorrelation. *NeuroImage* **220**, 117038 (2020).

29 Váša, F. *et al.* Adolescent tuning of association cortex in human structural brain networks. *Cerebral Cortex* **28**, 281-294 (2018).

30 Betzel, R. F. & Bassett, D. S. Specificity and robustness of long-distance connections in weighted, interareal connectomes. *Proceedings of the National Academy of Sciences*, 201720186, doi:10.1073/PNAS.1720186115 (2018).

31 Raj, A., Kuceyeski, A. & Weiner, M. A network diffusion model of disease progression in dementia. *Neuron* **73**, 1204-1215, doi:10.1016/j.neuron.2011.12.040 (2012).

32 Raj, A. *et al.* Network diffusion model of progression predicts longitudinal patterns of atrophy and metabolism in Alzheimer's disease. *Cell reports* **10**, 359-369 (2015).

33 Nichols, T. & Hayasaka, S. Controlling the familywise error rate in functional neuroimaging: a comparative review. *Statistical methods in medical research* **12**, 419-446 (2003).

34 Rubinov, M. & Sporns, O. Complex network measures of brain connectivity: uses and interpretations. *Neuroimage* **52**, 1059-1069 (2010).

35 Shafiei, G. *et al.* Spatial patterning of tissue volume loss in schizophrenia reflects brain network architecture. *Biological psychiatry* **87**, 727-735 (2020).
